# Genomic Variation Predicts Real-Time Δ^9^-tetrahydrocannabinol Response in Humans

**DOI:** 10.64898/2026.06.26.26356685

**Authors:** Uri Bright, Suhas Ganesh, Daniel F. Levey, Priya Gupta, the Yale THC Studies Consortium, Mohini Ranganathan, the IOP THC Studies Consortium, Robin M Murray, Marta DiForti, Paul Morrison, Deepak Cyril D’Souza, Joel Gelernter

## Abstract

**Background:** Cannabis is one of the most widely used psychoactive substances worldwide. Δ^9^-tetrahydrocannabinol (Δ^9^-THC) is the main contributor to cannabis-induced effects such as euphoria, anxiety, and psychotomimetic effects, and is metabolized by several hepatic enzymes, including CYP3A4. There are interindividual differences in how cannabis affects users, which have substantial genetic contributors.

**Methods:** We examined how real-time effects of Δ^9^-THC on psychotomimetic measures and on subjective effects of “high”, sadness and anxiety in 188 healthy volunteers in a laboratory infusion paradigm, relate to polygenic risk scores (PRS) for cannabis lifetime use (CanLU), cannabis use disorder (CanUD), and CYP3A4 expression.

**Results:** CYP3A4 expression PRS was significantly associated with Δ^9^-THC-induced psychotomimetic effects. Genetic liability to use and misuse cannabis is potentially associated with lower Δ^9^-THC-induced psychotomimetic symptoms. CanLU PRS nominally predicted enhanced Δ^9^-THC-induced “high”, while CanUD PRS predicted it to be lower.

**Conclusions:** Our findings suggest that genetic liability to produce more CYP3A4 enzyme may be associated with faster Δ^9^-THC degradation and the consequential diminution of the latter’s effects. Nominal effects suggest that aversive outcomes may reduce cannabis use and use disorder genetic liability, and that CanUD subjects may need higher Δ^9^-THC doses to experience euphoria (“high”). In total, this study provides novel insights regarding some of the specific genetic factors that influence interindividual variability in Δ^9^-THC effects, mainly via Δ^9^-THC metabolism.

## Introduction

Cannabis affects different people differently. These differences affect dependence risk, other adverse effects, and could plausibly also influence any therapeutic effects. Cannabis is among the most widely used psychoactive substances globally.(1, 2) Cannabis contains an array of bioactive compounds, including cannabinoids, terpenoids, and flavonoids. Δ^9^-tetrahydrocannabinol (Δ^9^-THC) is the principal psychoactive constituent and is primarily responsible for its intoxicating effects.

Typical acute effects of cannabis include euphoria, relaxation, altered perception, impaired cognition (particularly attention, memory, and executive function), tachycardia, dry mouth, conjunctival injection, increased appetite, dizziness or lightheadedness, and impaired psychomotor coordination and reaction time. With smoking, these effects generally emerge rapidly and last for a few hours.(3, 4)Cannabis can also be anxiogenic and induce panic, paranoia, or transient psychotic symptoms, effects that are also more likely to occur with products containing high amounts of THC, including cannabis concentrates, and products containing synthetic cannabinoids (e.g., “spice”). These effects are hard to quantify by retrospective self-report. Human laboratory studies provide a much more suitable context because of the ability to standardize dosing and to measure outcomes while they are being experienced.

Cannabis effects vary depending on the route of administration (e.g. smoking, eating),(5) potency (Δ^9^-THC content), the THC:CBD content, and context (social setting and expectancy).(6) Substantial inter- and intra-individual differences in the intoxicating effects of cannabis have been observed.(7–9)

Differences in Δ^9^-THC metabolism could also contribute to the variability in acute effects. Δ^9^-THC is primarily metabolized in the liver via cytochrome P450 enzymes, most notably CYP2C9, CYP2C19, and CYP3A4, although additional enzymes also contribute.(10–12) CYP3A enzymes play a central role in the oxidation of Δ^9^-THC to its psychoactive metabolite, 11-hydroxy-THC (11-OH-THC), and the subsequent metabolism of 11-OH-THC to the inactive metabolite 11-nor-9-carboxy-THC (COOH-THC).(10–13) The major Δ^9^-THC metabolites, 11-OH-THC and 11-COOH-THC, inhibit CYP2C9 but not CYP3A4, suggesting potential implications for inter-individual differences in THC pharmacokinetics and drug–drug interactions.(14)

Repeated cannabis use can lead to use disorder (CanUD), with approximately one in three regular users developing CanUD. Genome-wide association studies (GWAS) and post-GWAS analyses have demonstrated that CanUD is not only phenotypically associated with schizophrenia, but also shares moderate genetic liability with schizophrenia (genetic correlation rg ≈ 0.31–0.37).(15, 16) Genetic correlations between schizophrenia and lifetime cannabis use (CanLU) are rg ≈ 0.22–0.24.(15, 17, 18)

The goal of this investigation was to identify specific genetic influences on human response to Δ^9^-THC challenge, making use of data accumulated over decades at Yale University School of Medicine, U.S.A and at Kings College, London, U.K using similarly-designed human laboratory studies. We examined the relationship between polygenic risk for CanUD and CanLU, genetically predicted CYP3A4 expression, and the acute effects of Δ^9^-THC assessed in a highly controlled experimental human laboratory paradigm. Polygenic risk for CanUD and CanLU was estimated using previously published GWAS meta-analysis summary statistics for these phenotypes.(15, 18) Several variants may affect CYP3A4 function, including rs35599367, which maps to CYP3A4 and is associated with decreased CYP3A4 activity and expression.(19, 20) Other variants were reported to alter CYP3A4 function or cause a complete loss of function, but these reports are not consistent; in a study that focused on a variety of these variants, all had minimal effects on CYP3A4-substrate metabolism.(21) However, a GWAS of CYP3A4 protein levels demonstrated that genetic variation detected accounts for 16.5% of the variance.(22) Therefore, for this study, genetic prediction of CYP3A4 expression (pQTL) was based on GWAS summary statistics of blood protein expression generated as part of a large-scale investigation of genetic regulation of protein levels in participants from the Fenland study.(22) These data provide a comprehensive set of genetic markers enabling polygenic risk score (PRS) prediction of CYP3A4 expression as a complex trait.

## Methods and Materials

### Participants, Procedure, Materials

Over the past three decades, the Schizophrenia Neuropharmacology Research Group at Yale (SNRGY) has conducted a series of randomized, double-blind, placebo-controlled, crossover studies characterizing the acute effects of intravenous Δ^9^-THC in human participants. Data from these studies were anonymized and stored in a secure centralized database using a common experimental protocol. For the present analyses, demographic characteristics, cannabis exposure measures, experimental conditions, and behavioral and subjective response outcomes were extracted for participants who provided DNA samples. All primary studies were conducted under an Investigational New Drug application (IND #51671) at the Neurobiological Studies Unit of the VA Connecticut Healthcare System, West Haven, Connecticut, USA, and were approved by the institutional review boards of the Yale University School of Medicine and the VA Connecticut Healthcare System. All procedures complied with ethical standards for human subjects’ research. Phenotypically comparable data from studies conducted at the Institute of Psychiatry, Psychology and Neuroscience (IOPPN), London, England, were incorporated and approved by the King’s College London Research Ethics Committee (RESCMR-16/17-4163); all participants provided written informed consent and the study was conducted in compliance with the principles of Good Clinical Practice, the Declaration of Helsinki (1996).

Leveraging this uniquely large repository, one of the largest known collection of human laboratory model (HLM) studies of cannabinoid effects, we examined genetic determinants of inter-individual variability in acute responses to Δ^9^-THC. Data from SNRGY and IOPPN studies were harmonized to ensure comparability across sites and study protocols. Only measures with conceptual and methodological equivalence were included. Variables were mapped to common definitions, units, and coding schemes, and response measures were aligned by dose, timing relative to Δ^9^-THC administration, and assessment instrument. Site and study indicators were retained and modeled as covariates to account for residual methodological differences.

The analysis included Δ^9^-THC response data from healthy volunteers who received a total of 297 intravenous Δ^9^-THC infusions across studies conducted at Yale University and the IOPPN between 1997 and 2017. Detailed methods for participant screening, inclusion and exclusion criteria, recruitment, randomization, blinding, Δ^9^-THC administration, pre- and post-infusion assessments, and safety monitoring have been published previously.(23–29) All studies excluded cannabis-naïve individuals; several studies were designed to include participants with a broad range of prior cannabis exposure.(29) Participants were requested to abstain from drugs and medications for at least one week prior to each test day to minimize potential pharmacological interactions, with the exception of cannabis in studies that enrolled cannabis users.

Six of the 10 studies included in the present analysis investigated interactions between Δ^9^-THC and another drug such as cannabidiol, haloperidol,(24) iomazenil,(25) naltrexone,(26) tolcapone,(27) and physostigmine (unpublished). Only data from Δ^9^-THC-alone test days were included for the current analyses. **Table 1** presents a summary of the studies that were included in the current work.

**Table 1:** Summary of studies included in the current analysis.

| Study title | Sample size | $\Delta^9$-THC doses in mg/kg | Duration of infusion in minutes | $\Delta^9$-THC infusions with PANSS (n) |
| --- | --- | --- | --- | --- |
| The psychotomimetic effects of intravenous delta-9-tetrahydrocannabinol in healthy individuals: implications for psychosis (PMID: 15173844). | 44 | 0.035 and 0.07 | 2 to 5 | 87 |
| Effects of haloperidol on the behavioral, subjective, cognitive, motor, and neuroendocrine effects of Δ-9-tetrahydrocannabinol in humans (PMID: 18228005). | 31 | 0.0286 | 20 | 31 |
| Highs and lows of cannabinoid-dopamine interactions: effects of genetic variability and pharmacological modulation of catechol-O-methyl transferase on the acute response to delta-9-tetrahydrocannabinol in humans (PMID: 31187152). | 78 | 0.05 | 20 | 78 |
| Effects of Δ9-tetrahydrocannabinol in individuals with a familial vulnerability to alcoholism (PMID: 24424782). | 30 | 0.018 and 0.036 | 20 | 60 |
| Dose-related modulation of event-related potentials to novel and target stimuli by intravenous Δ9-THC in humans (PMID: 22334121). | 33 | 0.015 and 0.030 | 10 | 66 |
| GABA deficits enhance the psychotomimetic effects of Δ9-THC (PMID: 25728472). | 23 | 0.015 | 10 | 23 |
| Naltrexone does not attenuate the effects of intravenous Δ9-tetrahydrocannabinol in healthy humans (PMID: 22243563). | 6 | 0.025 | 20 | 6 |
| Interactions between physostigmine and THC (Unpublished) | 6 | 0.015 | 10 | 6 |
| Delta-9-Tetrahydrocannabinol, cannabidiol, and acute psychotomimetic states: a balancing act of the principal phyto-cannabinoids on human brain and behavior (PMID: 35319274). | 29 | 0.035 | 20 | 29 |
| The psychosis-like effects of $\Delta$(9)-tetrahydrocannabinol are associated with increased cortical noise in healthy humans (PMID: 25913109). | 7 | 0.015 and 0.030 | 10 | 14 |
| **Total** | **287<sup>#</sup>** |  |  | **400** |

As described in the parent studies,(24–27) Δ^9^-THC was administered intravenously into a rapidly flowing saline line, either by manual injection or infusion pump, over 2–5 minutes, 10 minutes, or 20 minutes. Δ^9^-THC doses ranged from 0.015 to 0.070 mg/kg, with some studies administering two different Δ^9^-THC doses to the same participants on separate test days.(28, 29)

Demographic variables were collected for all participants (**Table 2**). Lifetime and past 30-day cannabis exposure were assessed using the Scale for the Assessment of Lifetime Cannabis Use (SALCU),(30) and/or the Timeline Follow-Back (TLFB) method.(31) Current and lifetime psychiatric diagnoses, as well as alcohol and tobacco use, were assessed using the Structured Clinical Interview for DSM disorders (SCID).(32) The National Adult Reading Test (NART)(33) was administered to exclude participants with evidence of compromised intellectual functioning.

**Table 2:**
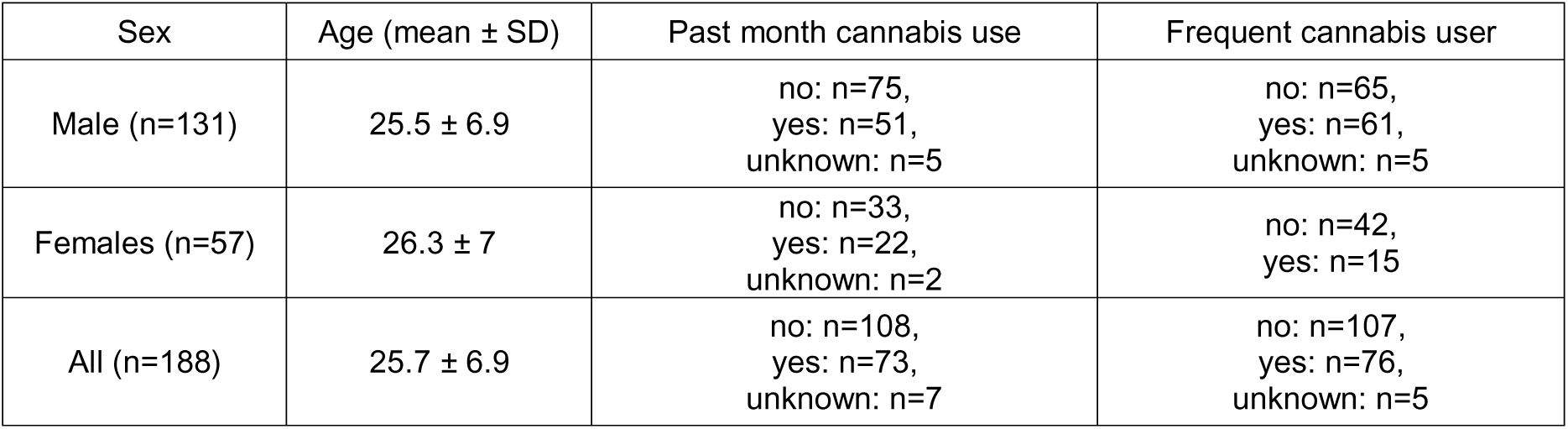
Demographics.

| Sex | Age (mean ± SD) | Past month cannabis use | Frequent cannabis user |
| --- | --- | --- | --- |
| Male (n=131) | 25.5 ± 6.9 | no: n=75,<br>yes: n=51,<br>unknown: n=5 | no: n=65,<br>yes: n=61,<br>unknown: n=5 |
| Females (n=57) | 26.3 ± 7 | no: n=33,<br>yes: n=22,<br>unknown: n=2 | no: n=42,<br>yes: n=15 |
| All (n=188) | 25.7 ± 6.9 | no: n=108,<br>yes: n=73,<br>unknown: n=7 | no: n=107,<br>yes: n=76,<br>unknown: n=5 |

The subjective reward-relevant effects of Δ^9^-THC including “high”, “anxiety,” and “sadness.” were measured by asking subjects to score on a 100-point visual analog scale (VAS) (0 = not at all to 100 = most of all) the intensity of their experience. The psychotomimetic effects of Δ^9^-THC were measured using the Positive and Negative Syndrome Scale (PANSS) which was modified to exclude items not appropriate for repeated assessment over short intervals (N4: Passive/apathetic social withdrawal; G16: Active social avoidance) [25]. The PANSS includes subscales for positive, negative, and general psychopathology symptoms. All the above assessments were administered prior to and after Δ^9^-THC infusion. For each outcome measure, the peak post-infusion score observed for each item was identified and summed to derive total and subscale scores, as appropriate. Baseline (pre-infusion) scores were subtracted from the corresponding post-infusion peak scores to generate change scores, which served as the primary indices of acute Δ^9^-THC response.

### Array genotyping and quality controls

Subjects were genotyped using the Illumina Infinium Global Diversity Array (KCL) or Illumina Infinium Psych Array (Illumina, San Diego CA USA) in the Gelernter lab (West Haven CT USA). Genotype information of 340 individuals was available. The data were subjected to standard quality control procedures using PLINK. Single nucleotide polymorphisms (SNPs) and samples with >10% missing data were excluded. SNPs with minor allele frequency <1% were excluded. There were no sex discrepancies detected and only autosomes were analyzed. The filtered dataset was evaluated for heterozygosity, and samples with heterozygosity rates falling outside ±3 standard deviations from the mean (n=3) were excluded. Thirty-one SNPs deviating from Hardy–Weinberg equilibrium expectations (<1e-06) were removed. The samples were assessed for relatedness, and eight pairs with a PI_HAT value > 0.2 (representing the proportion of alleles identical by descent; corresponding to second-degree relatedness or closer) were identified; one individual from each related pair was removed, leaving 322 unrelated samples as the final quality-controlled set.

The genetic ancestry of these samples was mapped using the 1000 genome reference panel.(34) 300 clustered with European ancestry (EUR) and were selected for PRS analysis. Genotypes for these were imputed using TOPMed.(35) Post-imputation, SNPs with an imputation quality score <0.8 were excluded, and the resulting dataset was subjected to the same quality control procedures described above. This yielded 300 samples with 4,776,236 SNPs each for PRS analysis.

### Final Sample

Of the genotyped samples, 188 EUR participants were left after removal of subjects due to missing phenotypic data. PANSS data were available for 188 participants; VAS results for anxiety and sadness exist for 130 participants, while VAS results of “high” exist for 112 participants. Of the 188 study participants, 131 were male, 76 were current cannabis users, and 73 had used cannabis in the prior month. The average age was 25.7 ± 6.9 years (**Table 2**).

### Functional Variant Associations

Though it did not survive quality controls procedures (imputation; --geno >0.28), we rescued for analysis rs35599367, a functional variant with known effects on CYP3A4 activity,(19) and tested its association with all the phenotypes that were included in this study. This test was conducted using PLINK 2,(36) using the same analytic parameters as for the GWAS.

### Polygenic Risk Score (PRS)

CanUD and CanLU PRS computation was conducted using our previously published summary statistics of GWAS meta-analyses of these traits.(15, 18) CYP3A4 expression PRS computation was conducted using GWAS summary statistics of CYP3A4 blood level, part of wide investigation of genetic regulation of protein expression in participants from the Fenland study (22). These summary statistics allow for genetic prediction of CYP3A4 in blood, including a comprehensive set of markers that allow for PRS prediction of CYP3A4 level as a complex genetic trait. PRS was calculated using PRSice-2,(37) with the 1000 genomes EUR data as a reference panel.(34)

These were used to evaluate polygenic association with positive symptoms, negative symptoms, general symptoms and total PANSS scores, and for sensations of high, anxiety and sadness measured using a visual analog scale (VAS) (as target traits), in participants who were administered Δ^9^-THC. We used age, sex, past month cannabis use (yes/no), frequent cannabis using (yes/no) and the first 5 genetic PCs as covariates. We performed p-value-informed clumping with a distance threshold of 250 kb and r^2^=0.1. For every pair of traits (base vs target), we assessed the best p-value threshold for inclusion of variants in the analysis (i.e., the p-value threshold that yields the most significant effect in the PRS analysis), out of nine possible cutoff thresholds: 5×10^-8^, 5X10^-7^, 5X10^-6^, 5X10^-5^, 5X10^-4^, 5X10^-3^, 5X10^-2^, 5X10^-1^ and 1. A 5% false discovery rate (FDR) Benjamini-Hochberg correction was applied for a total of 21 tests (3 base traits x 7 target traits). See **Figure 1** for a summary of the study design.

**Figure 1:**
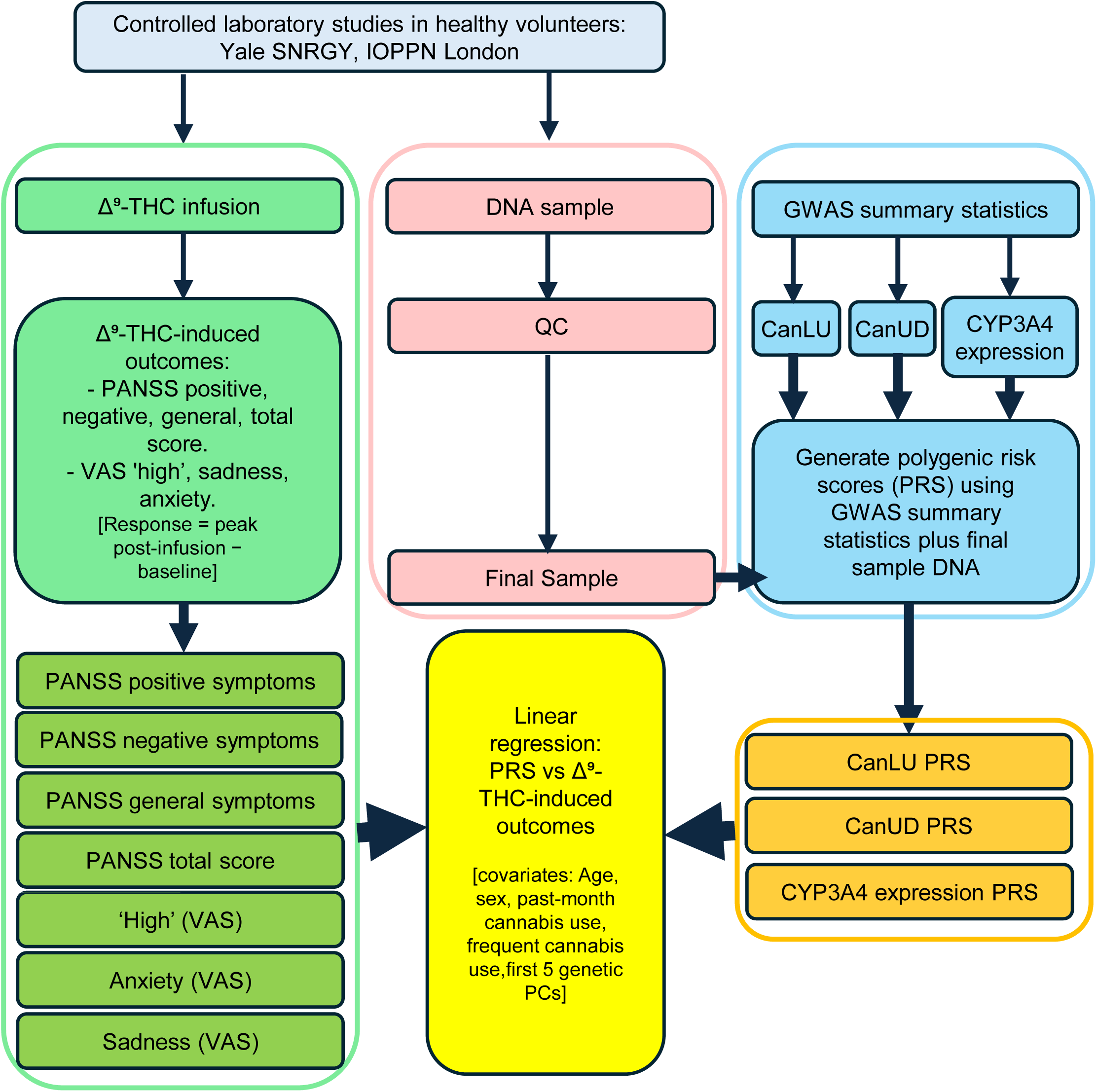
Study design. Polygenic risk scores for cannabis lifetime use, cannabis use disorder, and CYP3A4 expression were generated separately using GWAS summary statistics and participants’ DNA samples, then tested against seven Δ^9^-THC-induced outcomes across nine PRS thresholds using linear regression with covariates.

## Results

### Polygenic Risk Scores (PRS)

After correction for FDR, CYP3A4 expression PRS had significant effects on PANSS negative symptoms (r^2^=0.077, p=2.0×10^-4^), general symptoms (r^2^=0.05, p=3.5×10^-3^) and total symptoms (r^2^=0.055, p=2.1×10^-3^), with genetically-predicted CYP3A4 expression explaining 7.7%, 5.0% and 5.5% of the variance, respectively. In all these cases, the effect was negative, suggesting that higher CYP3A4 activity predicts lower psychotomimetic effects of Δ^9^-THC.

Several other tests had nominal significance (p<0.05) which did not survive FDR correction. At the nominal level, CYP3A4 expression predicted PANSS positive symptoms and anxiety; CanLU predicted PANSS positive symptoms, PANSS total score and VAS high; and CanUD predicted PANSS positive symptoms, PANSS general symptoms and VAS high. **Tables 3-5** present the summary of the results for the best-fit p-value threshold of each of the PRS analyses conducted. **Tables S1-3** and **Figures S1-S21** include the full PRS results for all analyses conducted in each of the p-value thresholds for SNP inclusion.

**Table 3:** Cannabis Lifetime Use PRS predictive value for psychotomimetic symptoms (PANSS) and subjective effects (VAS). The table presents the best-fit p-value threshold for SNP inclusion for each of the phenotypes. *suggestive statistical significance (p<0.05). [PANSS: positive and negative syndrome scale for schizophrenia; VAS: visual analog score].

| Phenotype | Threshold | PRS.R2 | Full.R2 | Null.R2 | Coefficient | SE | P | N SNPs |
| --- | --- | --- | --- | --- | --- | --- | --- | --- |
| PANSS Positive | $5.00 \times 10^{-6}$ | 0.035 | 0.110 | 0.078 | -460.3 | 186.9 | **0.015\*** | 87 |
| PANSS Negative | $5.00 \times 10^{-7}$ | 0.012 | 0.339 | 0.330 | -138.2 | 95.3 | 0.149 | 33 |
| PANSS General | $5.00 \times 10^{-6}$ | 0.017 | 0.257 | 0.244 | -520.1 | 307.2 | 0.092 | 87 |
| PANSS Total | $5.00 \times 10^{-6}$ | 0.028 | 0.286 | 0.265 | -1182.2 | 542.5 | **0.031\*** | 87 |
| VAS Anxiety | $5.00 \times 10^{-4}$ | 0.020 | 0.049 | 0.030 | 10752.5 | 7234.2 | 0.140 | 1033 |
| VAS High | $5.00 \times 10^{-8}$ | 0.048 | 0.111 | 0.066 | 1756.1 | 819.0 | **0.034\*** | 14 |
| VAS Sadness | $5.00 \times 10^{-8}$ | 0.022 | 0.104 | 0.084 | 496.6 | 318.6 | 0.122 | 14 |

**Table 4:**
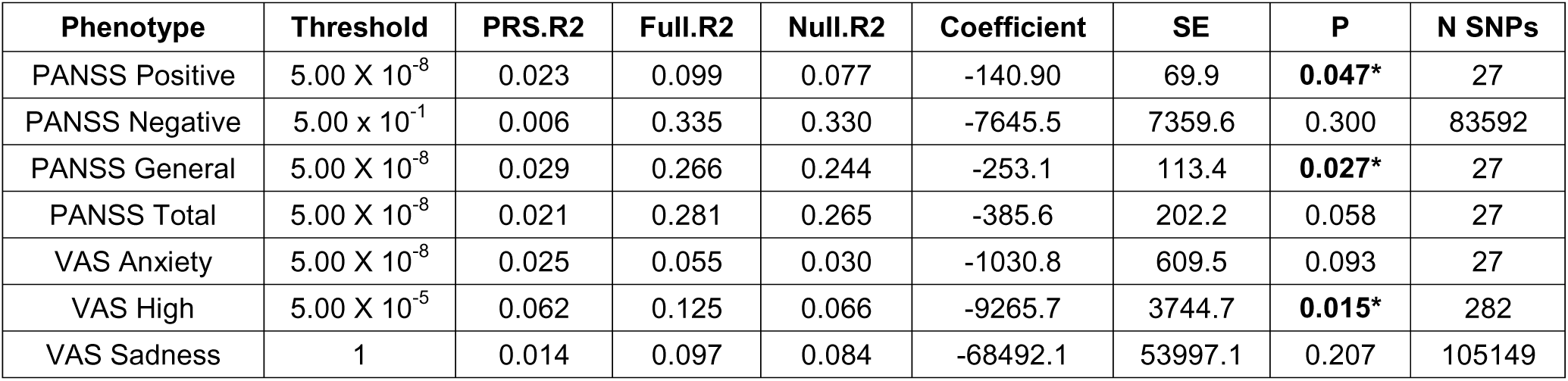
Cannabis Use Disorder PRS predictive value for psychotomimetic symptoms (PANSS) and subjective effects (VAS). The table presents the best-fit p-value threshold for SNP inclusion for each of the phenotypes. *suggestive statistical significance (p<0.05). [PANSS: positive and negative syndrome scale for schizophrenia; VAS: visual analog score].

| Phenotype | Threshold | PRS.R2 | Full.R2 | Null.R2 | Coefficient | SE | P | N SNPs |
| --- | --- | --- | --- | --- | --- | --- | --- | --- |
| PANSS Positive | $5.00 \times 10^{-8}$ | 0.023 | 0.099 | 0.077 | -140.90 | 69.9 | **0.047\*** | 27 |
| PANSS Negative | $5.00 \times 10^{-1}$ | 0.006 | 0.335 | 0.330 | -7645.5 | 7359.6 | 0.300 | 83592 |
| PANSS General | $5.00 \times 10^{-8}$ | 0.029 | 0.266 | 0.244 | -253.1 | 113.4 | **0.027\*** | 27 |
| PANSS Total | $5.00 \times 10^{-8}$ | 0.021 | 0.281 | 0.265 | -385.6 | 202.2 | 0.058 | 27 |
| VAS Anxiety | $5.00 \times 10^{-8}$ | 0.025 | 0.055 | 0.030 | -1030.8 | 609.5 | 0.093 | 27 |
| VAS High | $5.00 \times 10^{-5}$ | 0.062 | 0.125 | 0.066 | -9265.7 | 3744.7 | **0.015\*** | 282 |
| VAS Sadness | 1 | 0.014 | 0.097 | 0.084 | -68492.1 | 53997.1 | 0.207 | 105149 |

**Table 5:** CYP3A4 expression PRS predictive value for psychotomimetic symptoms (PANSS) and subjective effects (VAS). anxiety, high and sadness. The table presents the best-fit p-value threshold for SNP inclusion for each of the phenotypes. *suggestive statistical significance (p<0.05), **FDR statistical significance (p<0.007). [PANSS: positive and negative syndrome scale for schizophrenia; VAS: visual analog score].

| Phenotype | Threshold | PRS.R2 | Full.R2 | Null.R2 | Coefficient | SE | P | N SNPs |
| --- | --- | --- | --- | --- | --- | --- | --- | --- |
| PANSS Positive | $5.00 \times 10^{-8}$ | 0.025 | 0.100 | 0.077 | -62.4 | 30.3 | 0.041* | 13 |
| **PANSS Negative** | $5.00 \times 10^{-7}$ | 0.077 | 0.382 | 0.330 | -108.4 | 29.0 | **0.0002\*\*** | **17** |
| **PANSS General** | 1 | 0.050 | 0.282 | 0.244 | -55389.5 | 18687.1 | **0.004\*\*** | **103922** |
| **PANSS Total** | $5.00 \times 10^{-8}$ | 0.055 | 0.306 | 0.265 | -269.1 | 86.2 | **0.0021\*\*** | **13** |
| VAS Anxiety | $5.00 \times 10^{-8}$ | 0.038 | 0.068 | 0.030 | -555.0 | 264.8 | 0.038* | 13 |
| VAS High | $5.00 \times 10^{-4}$ | 0.016 | 0.081 | 0.067 | 4249.3 | 3488.3 | 0.226 | 355 |
| VAS Sadness | 1 | 0.025 | 0.106 | 0.084 | -95313.8 | 57229.1 | 0.098 | 103922 |

### Functional Variant Associations

*CYP3A4*\*rs35599367, a reportedly functional variant, was observed in six heterozygotes among the available 99 imputed samples (lower than the full sample because of incomplete imputation). There was no statistically significant association for this variant (p<0.05) with any of the cannabis-induced effects measured for which a test could be conducted (for some of the traits, the variant count was too low to analyze; **Table S4**).

## Discussion

This is the first study, to our knowledge, linking polygenic liability to experimentally measured responses to Δ^9^-THC in humans. The evidence provided links genetic variation with acute Δ^9^-THC effects and, by extension, real-world cannabis experiences. Using PRS derived from our GWAS of cannabis-related traits and published GWAS of CYP3A4 expression, we evaluated whether PRS for CanLU, CanUD, and pQTL-based CYP3A4 blood level expression predicts Δ^9^-THC–induced psychotomimetic effects and subjective effects in a controlled human laboratory paradigm. The strongest findings involved CYP3A4. Polygenic risk for increased CYP3A4 expression, a key determinant of Δ^9^-THC metabolism,(10–12) was associated with attenuated Δ^9^-THC–induced negative and general symptoms and total PANSS scores and with nomina associations for lower anxiety and positive symptoms. Mechanistically, greater genetically driven CYP3A4 expression is plausibly associated with faster clearance of parent Δ^9^-THC, lower circulating levels, and reduced central effects. These results provide evidence that an important factor influencing pharmacokinetic variation i.e., CYP3A4 expression shapes the effects of Δ^9^-THC.

In addition, PRS for CanLU and CanUD showed nominal associations with Δ^9^-THC–induced effects. Higher genetic liability for cannabis use and use disorder was associated with attenuated psychotomimetic responses, including lower PANSS positive and total scores. Although these findings did not survive correction for multiple comparisons and should be interpreted cautiously, they suggest that genetic liability for cannabis exposure and misuse may reflect differences in acute sensitivity to Δ^9^-THC. Higher CYP3A4 expression PRS was associated with significantly lower Δ^9^-THC–induced psychotomimetic symptoms, including negative and general symptoms and total PANSS score, with nominal associations for positive symptoms and anxiety. CYP3A4 plays a central role in Δ^9^-THC metabolism,(10–12) and greater genetically driven expression is plausibly associated with faster clearance of parent Δ^9^-THC, reduced circulating levels, and lower likelihood of psychotomimetic effects. We made these observations based on genomewide CYP3A4 protein level data. There is a variant, rs35599367 (MAF in EUR subjects in the 1000G reference panel = 0.0497) that affects function (as opposed to protein level) of CYP3A4. We rescued limited data for this variant from available GWAS array data, and it was not associated to any Δ^9^-THC outcomes. This result could reflect the low imputation quality or a true null effect. Genomewide PRS data proved to be a stronger approach.

For the CanLU and CanUD PRS, nominal associations (p < 0.05, not surviving FDR correction) with *lower* Δ^9^-THC–induced psychotomimetic symptoms (PANSS) were observed. Specifically, higher CanLU PRS was nominally associated with lower positive symptom scores on the PANSS and with a lower total PANSS score, whereas higher CanUD PRS was nominally associated with lower positive and general psychotomimetic symptoms. Although these findings should be interpreted cautiously, they suggest that greater genetic liability for cannabis use and misuse may be linked to attenuated acute Δ^9^-THC-induced psychotomimetic effects.

One plausible interpretation is that heightened sensitivity to aversive psychotomimetic effects may deter continued cannabis use and, by extension, liability for CanUD. In contrast, individuals with fewer adverse responses may be more likely to continue use and develop problematic patterns. This interpretation parallels findings in alcohol research, where blunted sensitivity to aversive effects is associated with increased risk for alcohol use disorder.(38) Within this framework, individuals with lower genetic liability for CanUD may be more susceptible to aversive psychotomimetic effects of acute Δ^9^-THC, which could deter continued use. Conversely, individuals with higher CanUD PRS may experience fewer acute psychotomimetic effects, facilitating continued use and increasing risk for disorder. This negative association between cannabis use traits and psychotomimetic symptoms coexists with epidemiological evidence showing higher prevalences of CanLU and CanUD among individuals with a history of psychosis,(39) as well as moderate positive genetic correlations between schizophrenia and CanLU (r_g_=0.22) and CanUD (r_g_=0.37).(15, 18) Our human laboratory sample excluded participants with a history of CanUD while requiring lifetime cannabis exposure, a design that likely restricted the range of CanUD genetic liability and may have accentuated inverse associations between CanUD PRS and acute Δ^9^-THC-induced psychotomimetic effects.

CanLU and CanUD PRS showed opposite nominal associations with the subjective sensation of being “high”. Higher CanLU PRS was associated with greater perceived high, whereas higher CanUD PRS was associated with a blunted high. This dissociation suggests that genetic liability for initiation versus problematic use may differentially influence sensitivity to rewarding versus aversive effects. In contrast to psychotomimetic effects, the experience of feeling high is typically perceived as rewarding and may be a primary driver of recreational cannabis use initiation (i.e., CanLU). Among individuals with higher liability for CanUD, reduced sensitivity to the rewarding effects of Δ^9^-THC may promote escalation of use to achieve desired intoxication, consistent with models of tolerance and reinforcement, and increasing risk for dependence.(40) This pattern is consistent with the possibility that pharmacokinetic factors such as faster Δ^9^-THC metabolism contribute to diminished rewarding effects, leading users to consume larger amounts of cannabis to obtain desired effects and thereby elevating CanUD risk. Similar patterns have been described in alcohol research, where reduced subjective intoxication is associated with increased consumption and risk for dependence.(41, 42)

The CYP3A4 blood protein-level GWAS underlying this PRS(22) identified genome-wide significant loci that do not map directly to the CYP3A4 gene, suggesting that regulation of CYP3A4 expression involves distal or trans-acting mechanisms. This further supports the use of polygenic approaches to capture biologically meaningful variation in drug metabolism that is not fully explained by known functional variants.

Our findings underscore the importance of specific genetic influences on both inferred pharmacokinetic and subjective responses to Δ^9^-THC. Associations that survived correction for multiple testing were confined to CYP3A4 expression, highlighting metabolism as a key determinant of acute response. Nominal associations with CanLU and CanUD PRS suggest broader genetic contributions beyond metabolism alone, but require replication in larger samples.

Identification of genetic factors influencing Δ^9^-THC response has potential clinical and public health relevance. Individuals with reduced capacity to metabolize Δ^9^-THC may be at elevated risk for psychotomimetic or anxiety-related effects, particularly with high Δ^9^-THC content products that are currently available on the market. Genetic or pharmacologic markers of Δ^9^-THC metabolism could eventually inform risk stratification, prevention efforts, or personalized guidance toward lower-Δ^9^-THC formulations in in both recreational and medical cannabis contexts.

Several limitations warrant consideration. Participants with a history of psychosis or CanUD were excluded. Individuals who previously had strong adverse cannabis reactions may have been less likely to enroll, potentially limiting generalizability. In addition, there were differences in protocols and Δ^9^-THC dosages across the different studies. Available GWAS data did not permit evaluation of other relevant metabolic enzymes such as CYP2C9 or CYP2C19, or of rare CYP3A4 variants presumed to affect enzyme function. The sample size precluded sex-stratified analyses; there are sex differences in both Δ^9^-THC effects(43) and CYP3A4 expression.(44) Finally, CYP3A4 expression is influenced by environmental factors, including exposure to other medications metabolized by CYP3A4.(45–48) Concomitant medication use was not controlled, and non-genetic variation in enzyme expression contributed to observed effects. However, most participants were healthy and were not taking concomitant medications, and any such effects would have added noise to our results, more likely leading to reduced observed genomic effects rather than false positives.

In conclusion, genetically predicted variation in CYP3A4 expression contributes to individual differences in psychotomimetic and anxiety-related responses to Δ^9^-THC. This supports that genetic factors influencing the metabolism of Δ^9^-THC may shape acute psychiatric sensitivity to cannabis. Nominal associations between cannabis use liability PRS and subjective THC effects suggest additional genetic mechanisms influencing risk and tolerance. Larger studies incorporating additional metabolic pathways, sex-specific effects, and longitudinal outcomes will be critical to fully elucidate the genetic architecture of cannabis response.

## Data Availability

All the phenotypic data used for this study is presented in the methods section, or in previously published studies referred to in the methods section. Full PRS results are available in supplementary files.

## Acknowledgments

We thank Ann Marrie Lacobelle for excellent laboratory assistance in the Gelernter genetics Lab, and Angelina Genovese RN and Elizabeth O’Donnell RN for their excellent nursing support for the Δ^9^-THC studies. We thank Andrew Sewell, MD, PhD, Patrick Skosnik, PhD (deceased), Rajiv Radhakrishnan, MD, PhD, and Ashley Schnakenberg-Martin, PhD from The Yale THC Studies Consortium. This research was supported by NIH grants R01DA058862 (J.G., D.C.D), R01DA054869 (J.G.), R01 DA12382 (DCD), R21DA030696 (MR), R21MH086769 (DCD), R21DA020750 (DCD), R21AA16311 (DCD); by funding from the Department of Veterans Affairs Office of Research and Development, USVA grant I01CX001849 (JG); by funding from the UK Medical Research Council (MRC), reference MR/T007818/1 (MDF); by funding of EU Youth-Gems, Psychiatry Research Trust, and Leverhulme Charitable Trust (RMM). SG is currently supported by an MQ: Transforming Mental Health fellowship MQF:22_20. Infrastructure support was provided by the National Center for PTSD, West Haven, CT and the Schizophrenia Neuropharmacology Research Group at Yale (SNRGY).

## Author Contribution

UB conducted collected and analyzed genotype data including the cannabis lifetime use GWAS and PRS computations, led the data analysis and drafted the article; SG conducted analysis of the phenotypic data, and provided a critical review of the manuscript. DFL conducted data analysis, performed the cannabis use disorder GWAS, and provided critical review of the manuscript; PG conducted data analysis, quality control of the novel microarray data, and provided critical review of the manuscript; MR collected the phenotypic data and conceived some of the human laboratory studies; RMM conceived some of the human laboratory studies, supervised the KCL work and provided a critical review of the manuscript; MDF supervised the KCL work and provided a critical review of the manuscript; PM collected the KCL phenotypic data and conceived some of the human laboratory studies; DCD conceived the study, drafted the article, and supervised the work; JG conceived the study, drafted the article, and supervised the work.

