## Supplementary Figures for "Genomic Variation Predicts Real-Time Δ^9^-tetrahydrocannabinol Response in Humans"

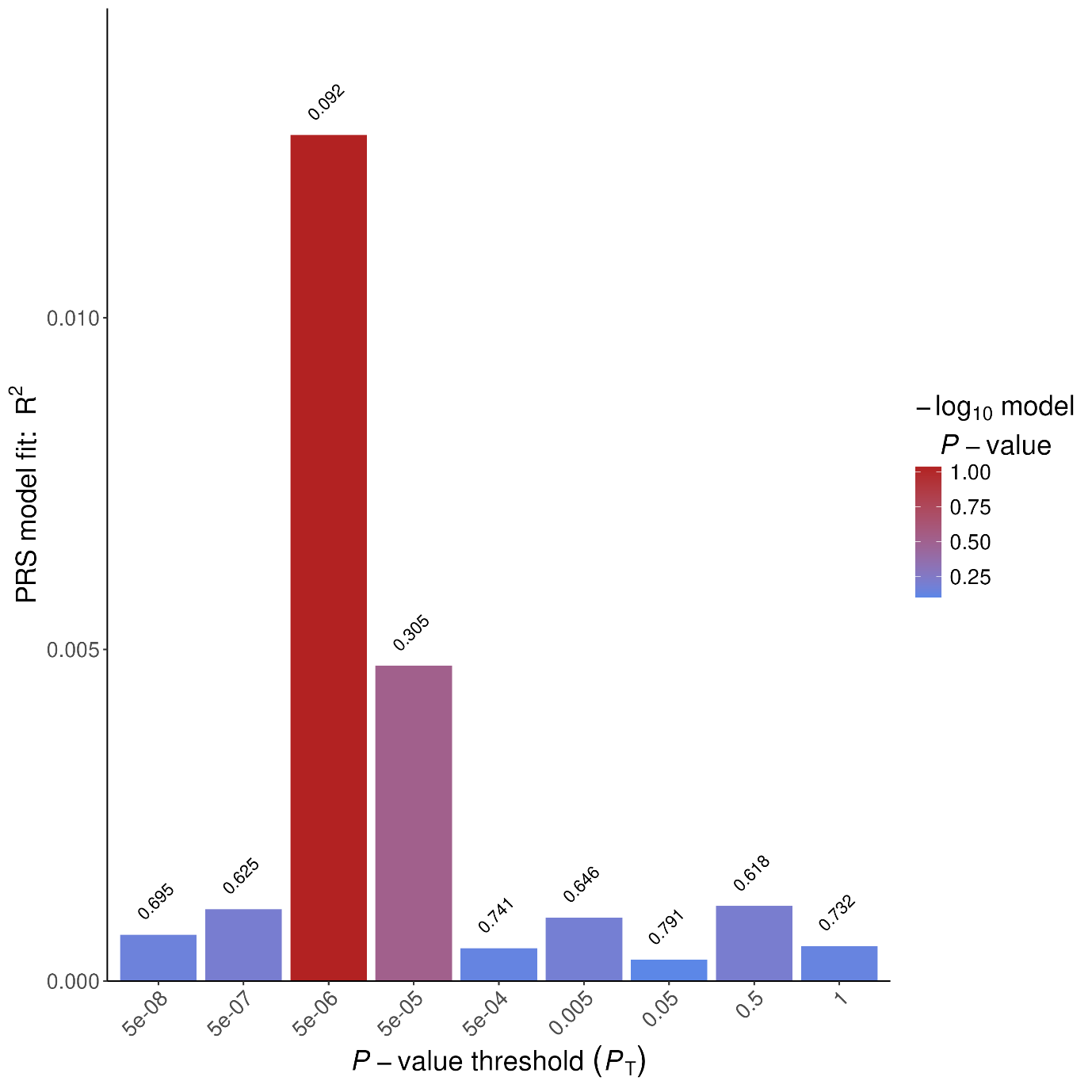

**Supplementary Figure S1: Prediction of Δ-9-THC-induced PANSS general symptoms by PRS of cannabis lifetime use across nine p-value thresholds for SNP inclusion**

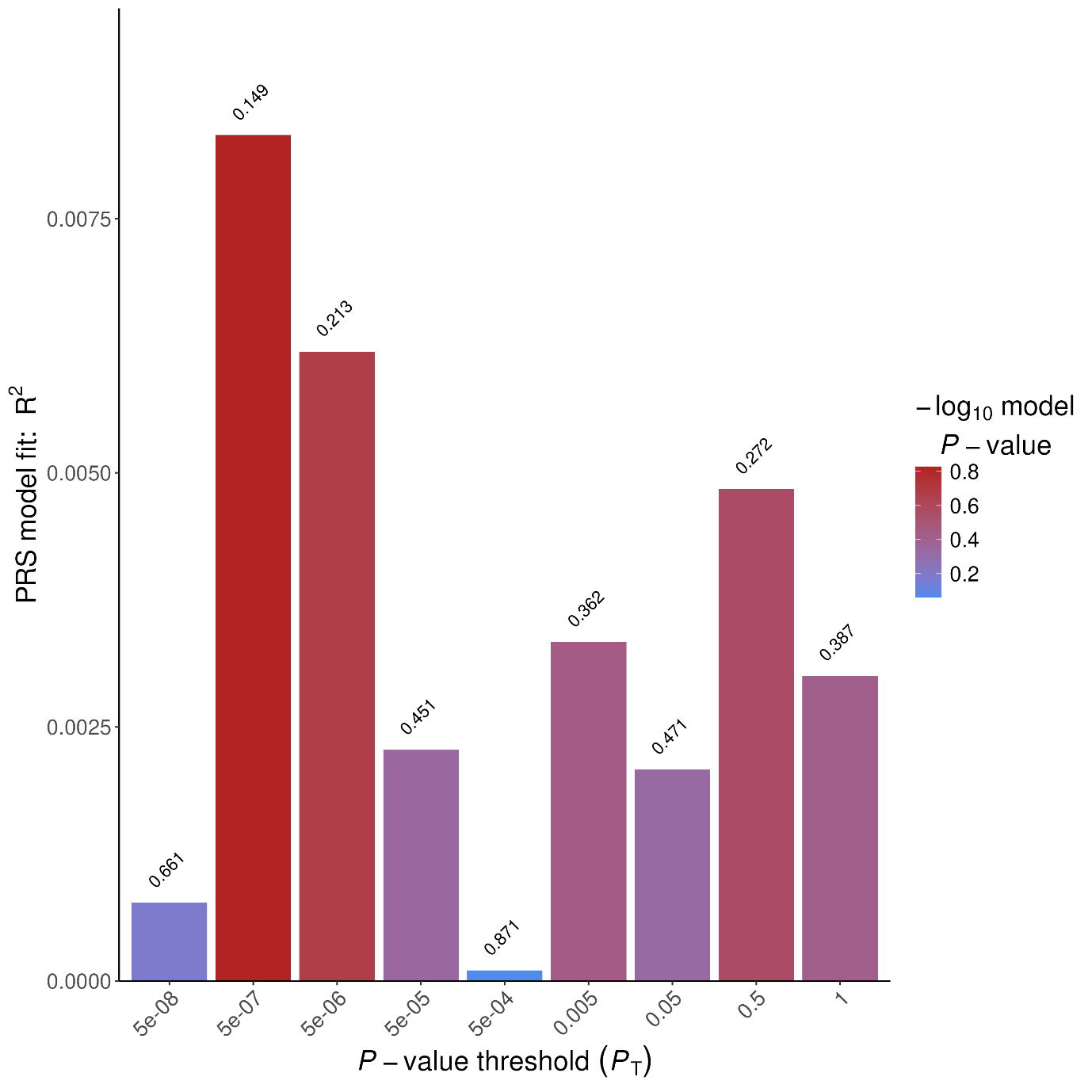

**Supplementary Figure S2: Prediction of Δ-9-THC-induced PANSS negative symptoms by PRS of cannabis lifetime use across nine p-value thresholds for SNP inclusion**

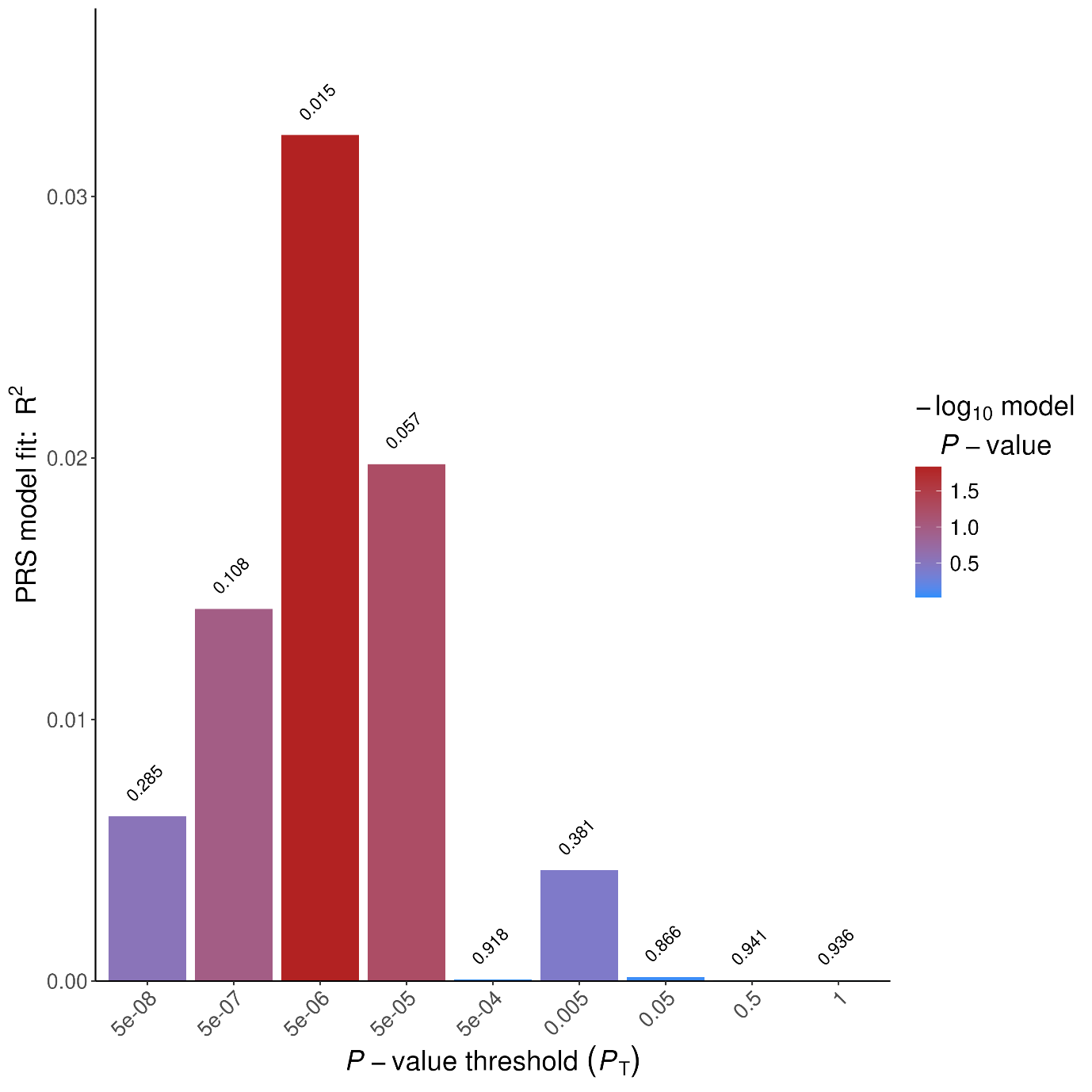

**Supplementary Figure S3: Prediction of Δ-9-THC-induced PANSS positive symptoms by PRS of cannabis lifetime use across nine p-value thresholds for SNP inclusion**

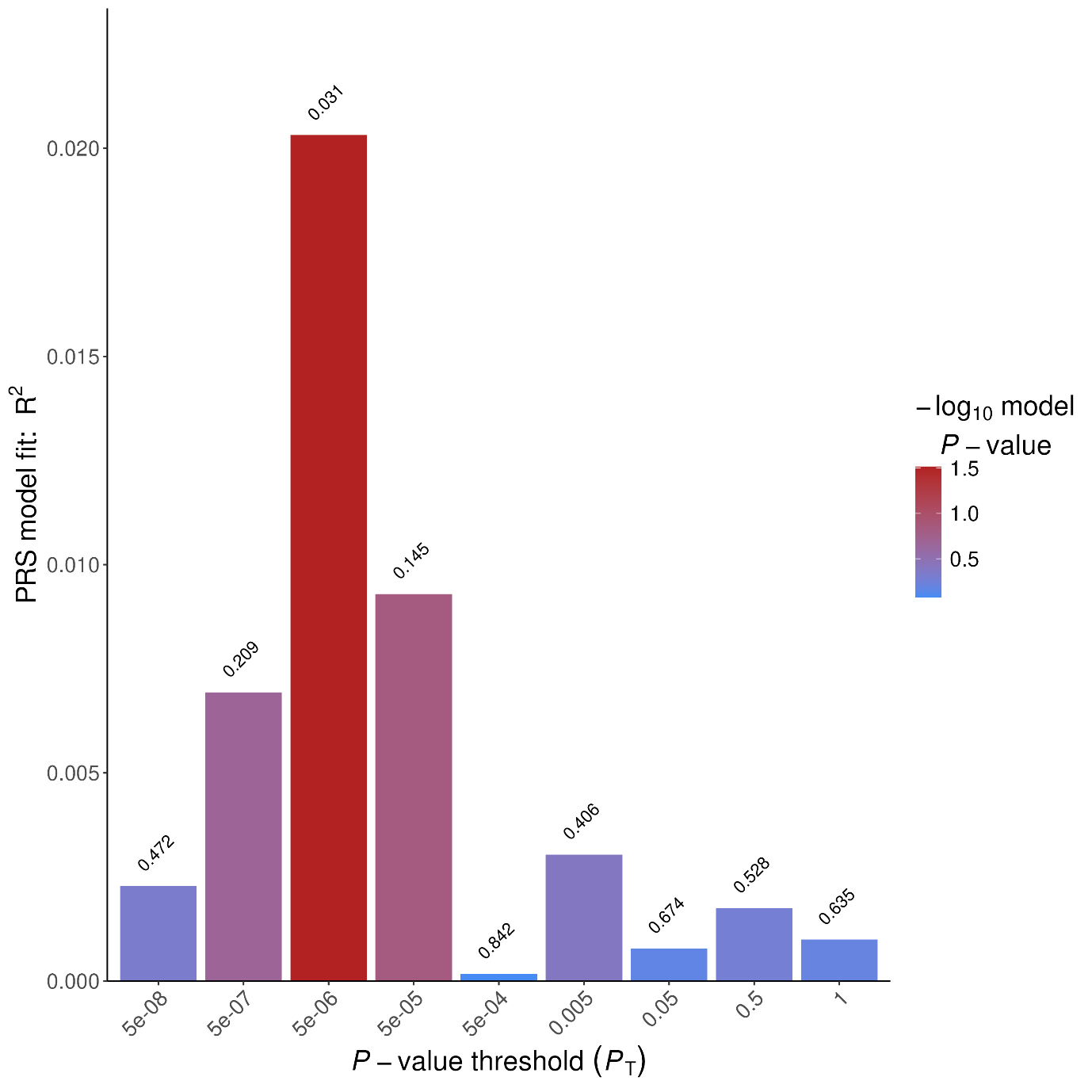

**Supplementary Figure S4: Prediction of Δ-9-THC-induced PANSS total score by PRS of cannabis lifetime use across nine p-value thresholds for SNP inclusion**

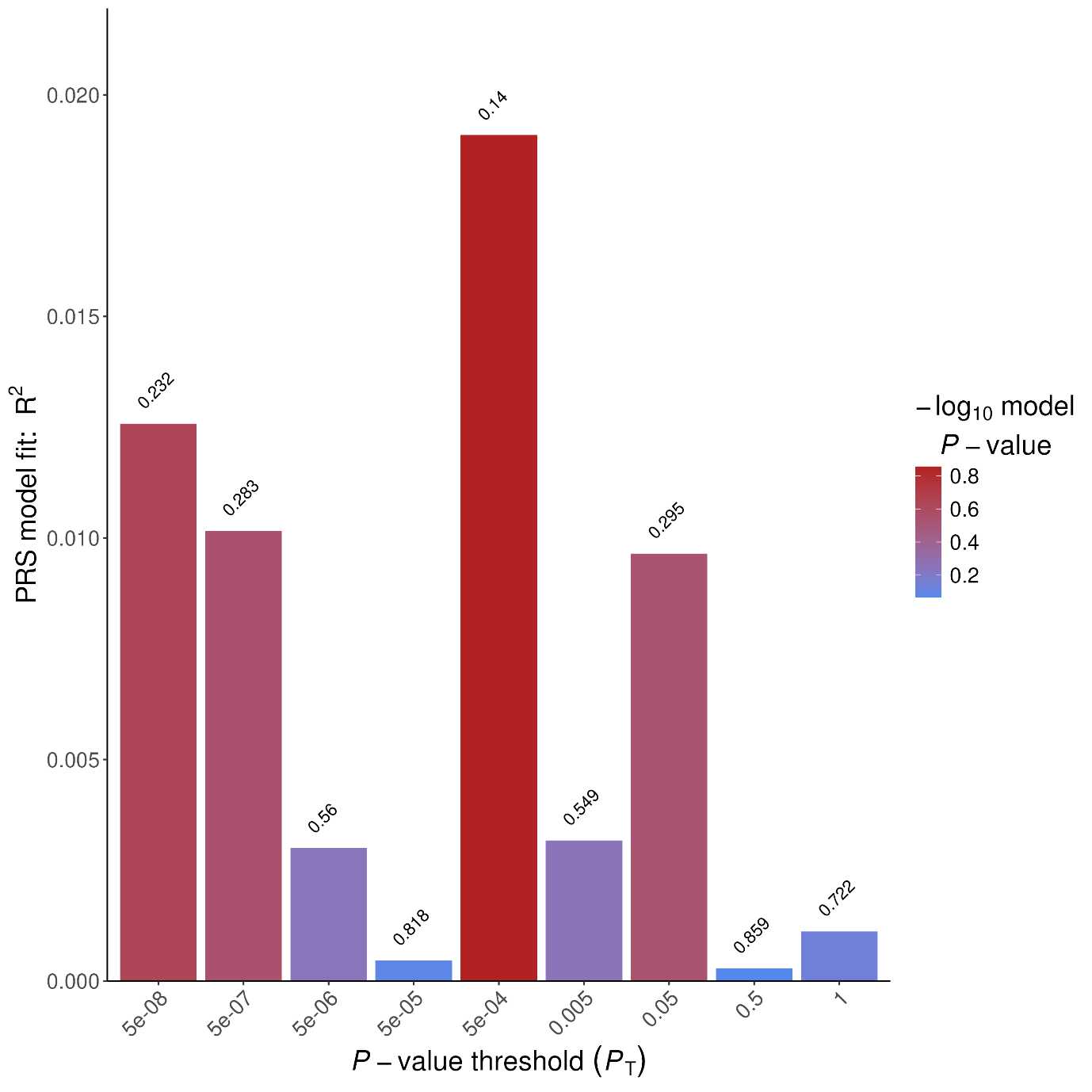

**Supplementary Figure S5: Prediction of Δ-9-THC-induced anxiety by PRS of cannabis lifetime use across nine p-value thresholds for SNP inclusion**

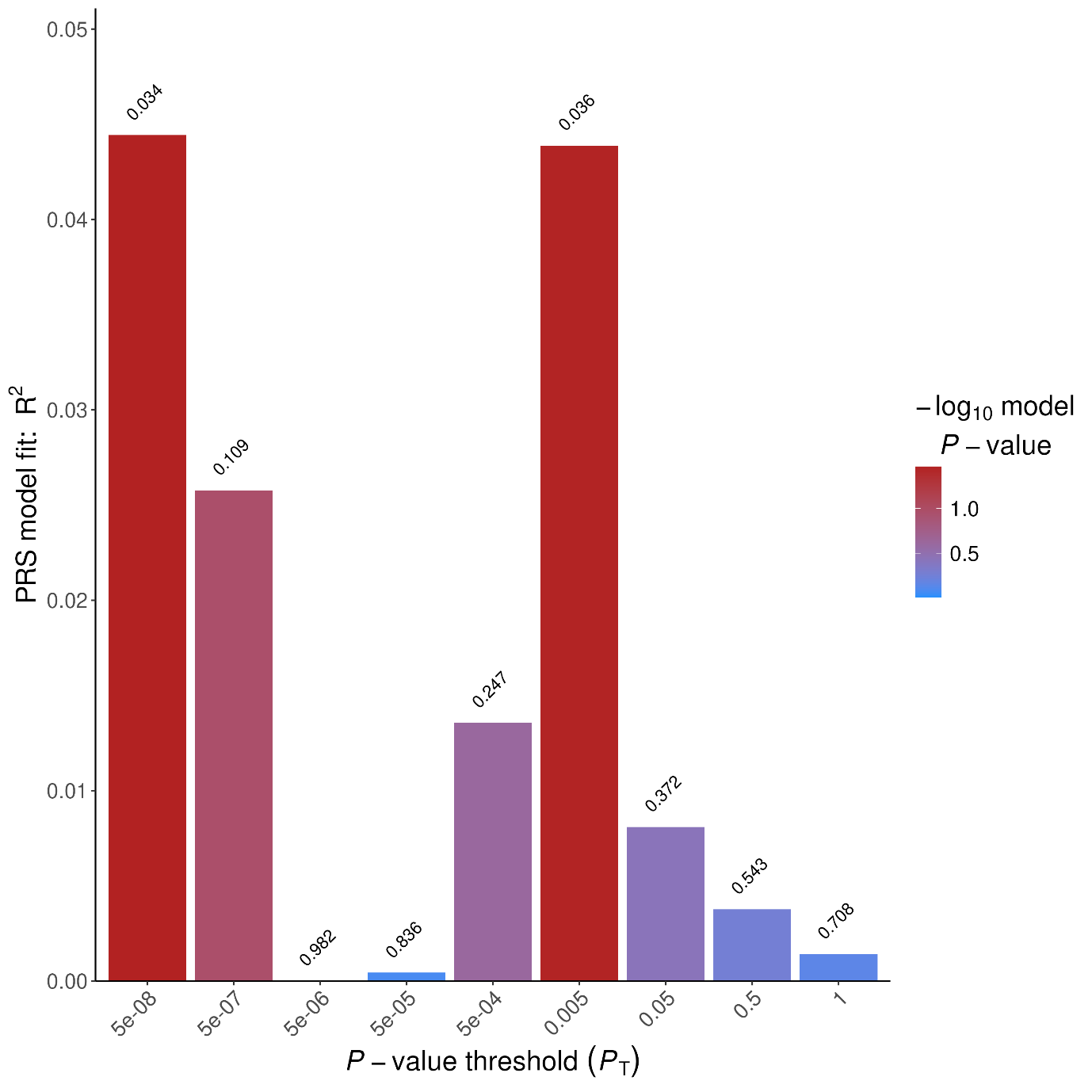

**Supplementary Figure S6: Prediction of Δ-9-THC-induced “high” by PRS of cannabis lifetime use across nine p-value thresholds for SNP inclusion**

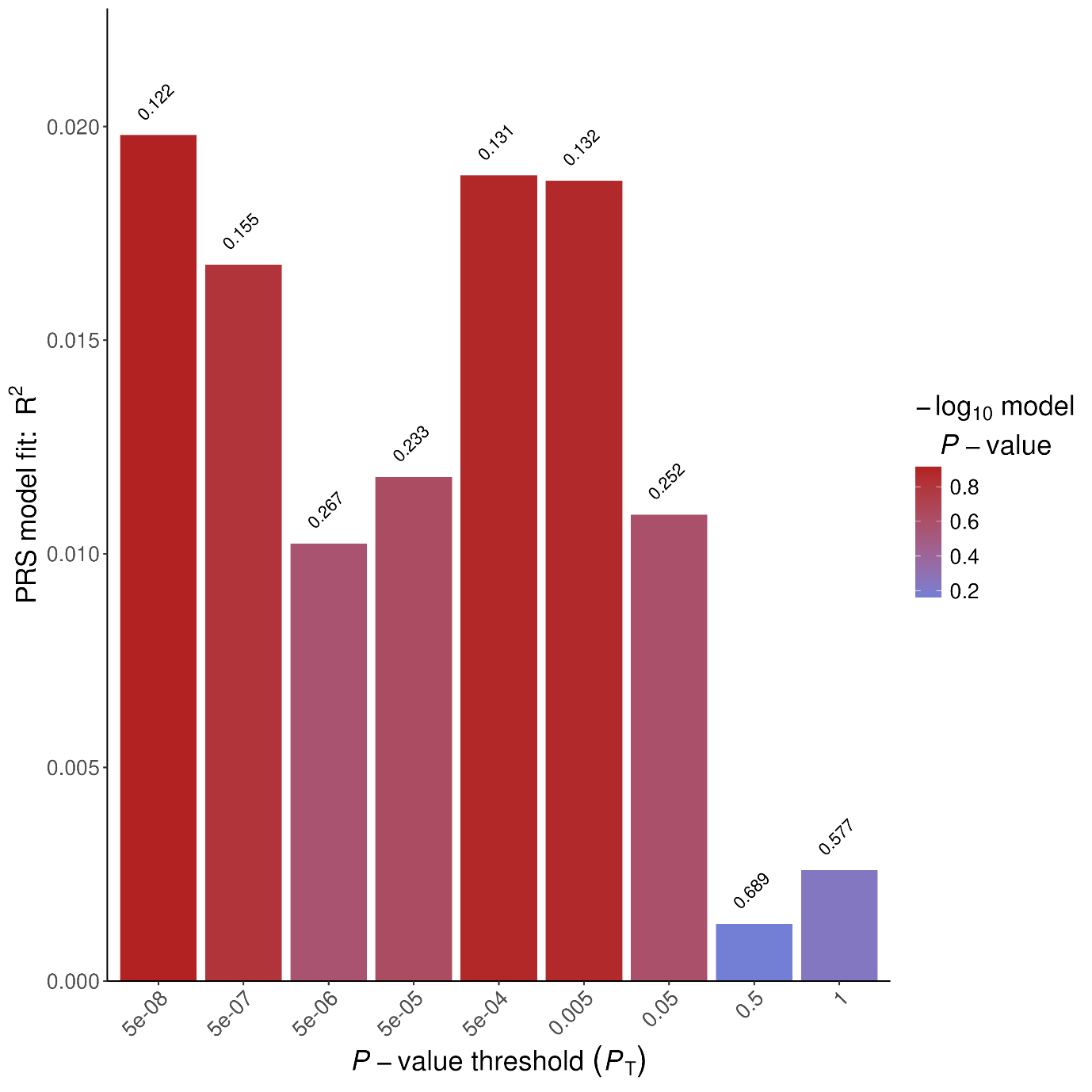

**Supplementary Figure S7: Prediction of Δ-9-THC-induced sadness by PRS of cannabis lifetime use across nine p-value thresholds for SNP inclusion**

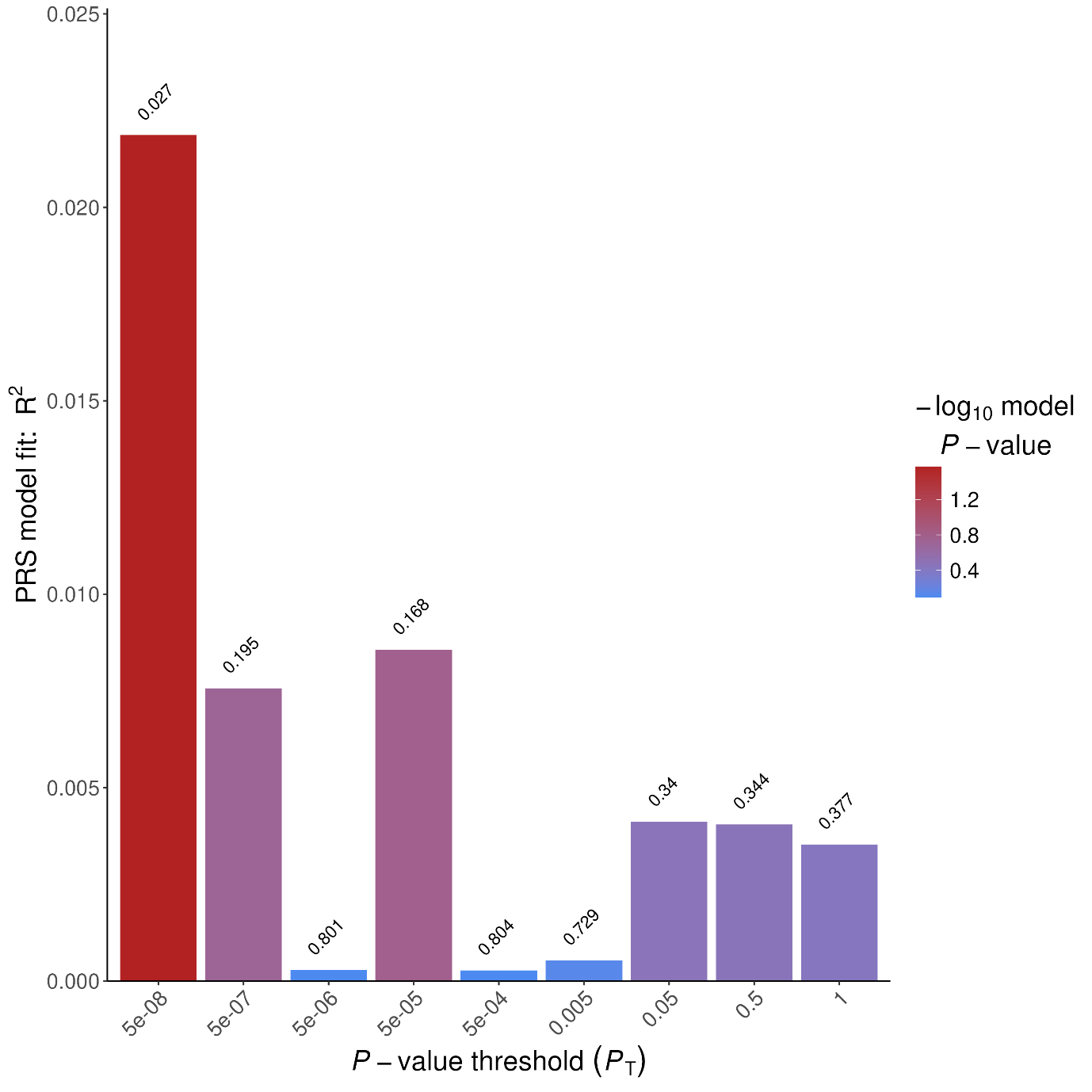

**Supplementary Figure S8: Prediction of Δ-9-THC-induced PANSS general symptoms by PRS of cannabis use disorder across nine p-value thresholds for SNP inclusion**

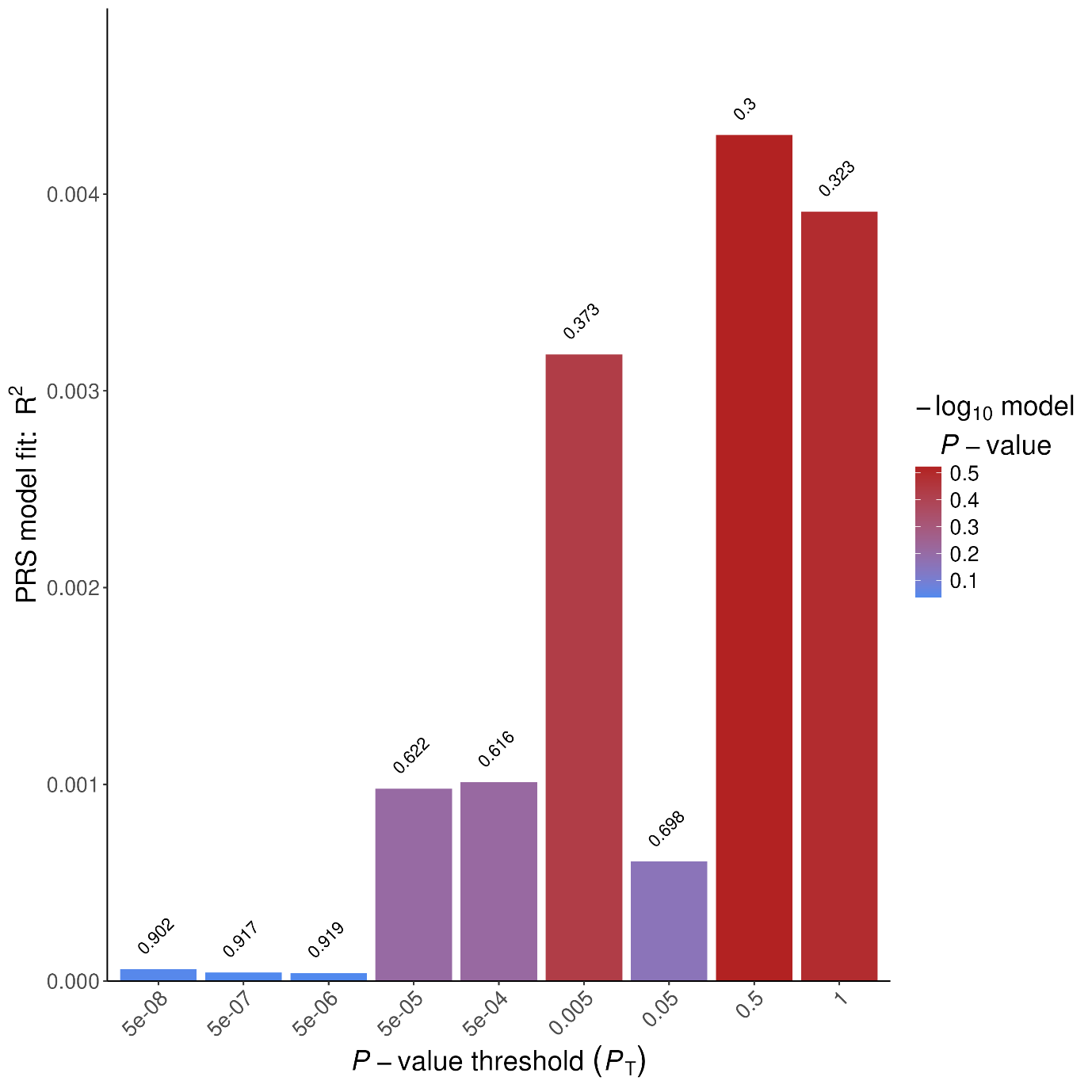

**Supplementary Figure S9: Prediction of Δ-9-THC-induced PANSS negative symptoms by PRS of cannabis use disorder across nine p-value thresholds for SNP inclusion**

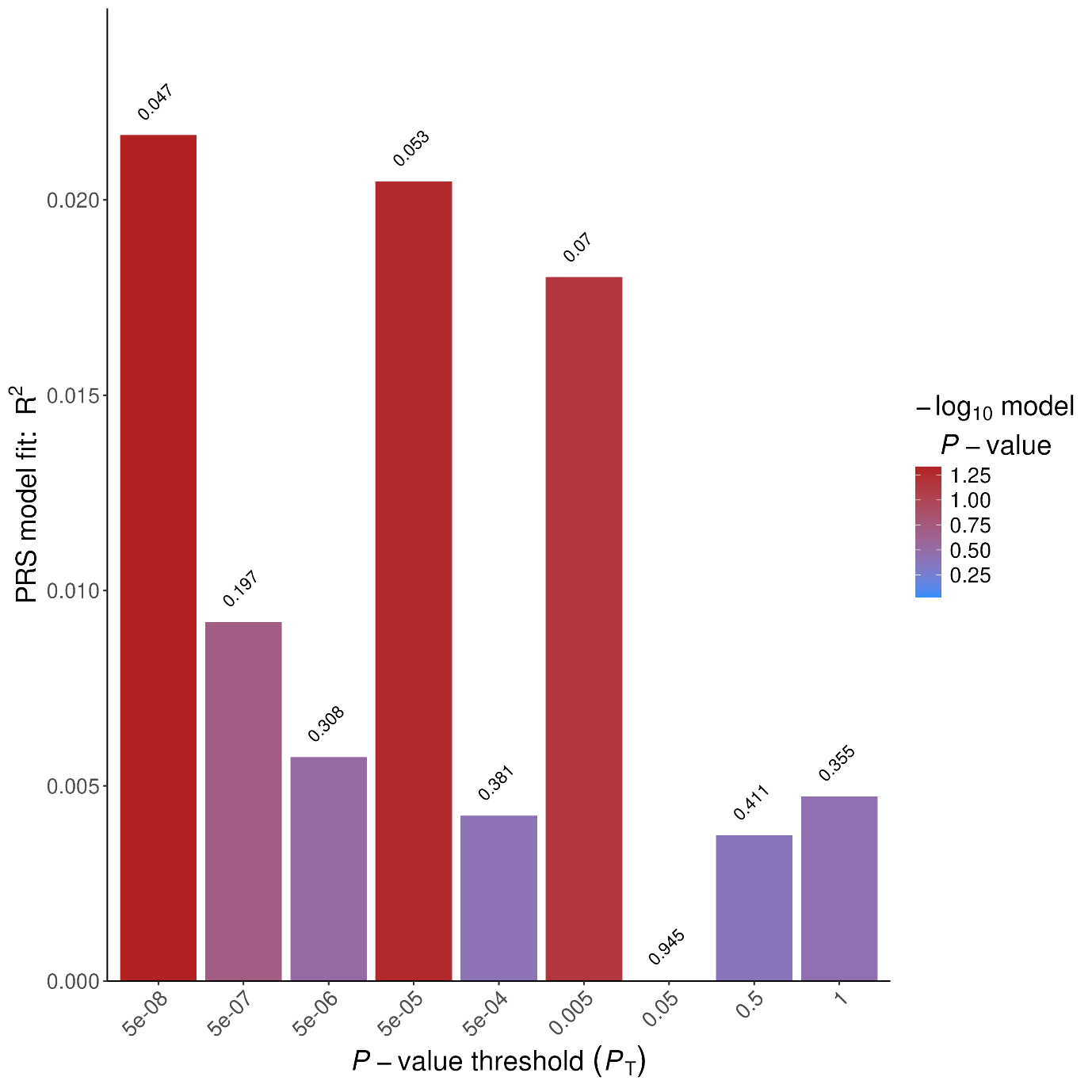

**Supplementary Figure S10: Prediction of Δ-9-THC-induced PANSS positive symptoms by PRS of cannabis use disorder across nine p-value thresholds for SNP inclusion**

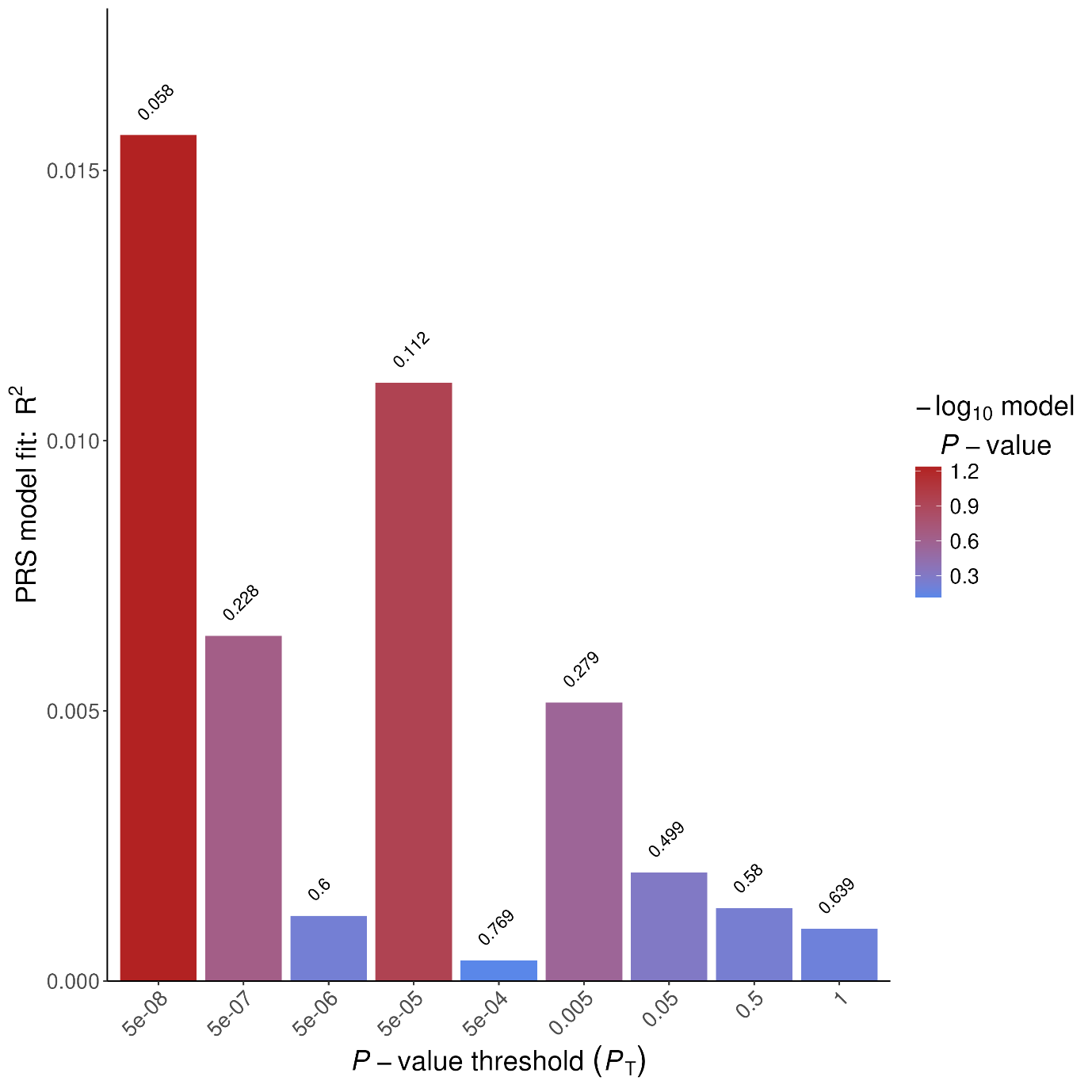

**Supplementary Figure S11: Prediction of Δ-9-THC-induced PANSS total score by PRS of cannabis use disorder across nine p-value thresholds for SNP inclusion**

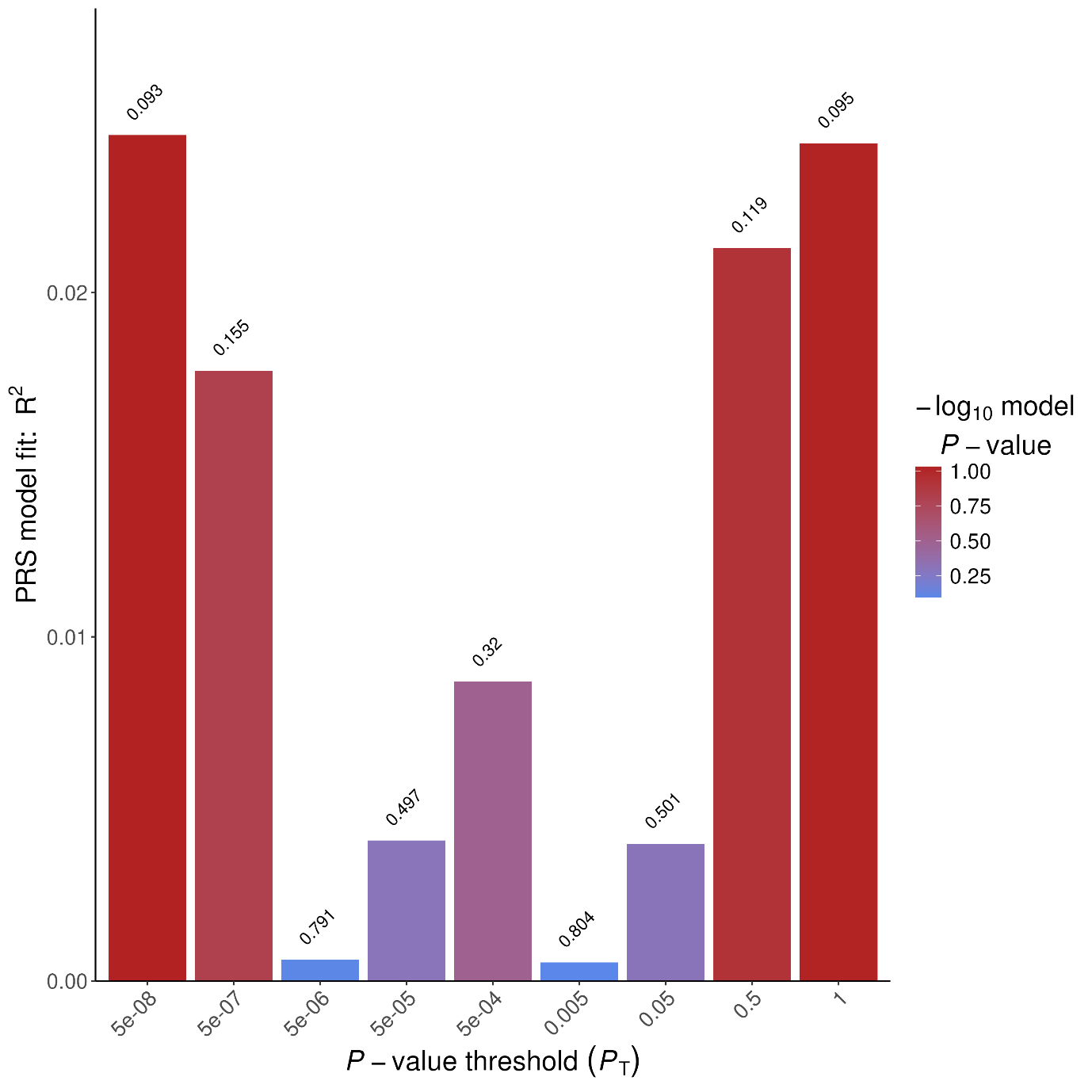

**Supplementary Figure S12: Prediction of Δ-9-THC-induced anxiety by PRS of cannabis use disorder across nine p-value thresholds for SNP inclusion**

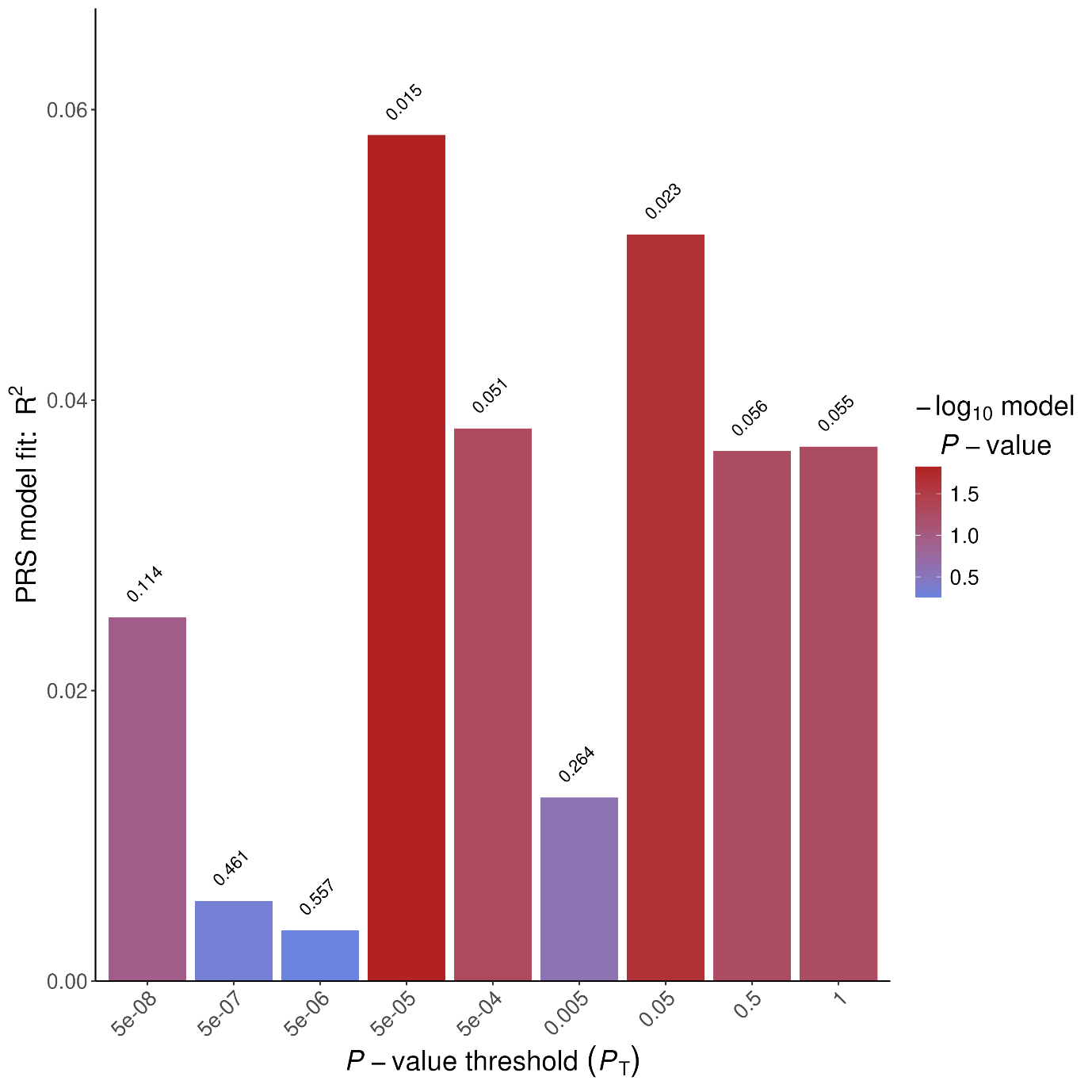

**Supplementary Figure S13: Prediction of Δ-9-THC-induced “high” by PRS of cannabis use disorder across nine p-value thresholds for SNP inclusion**

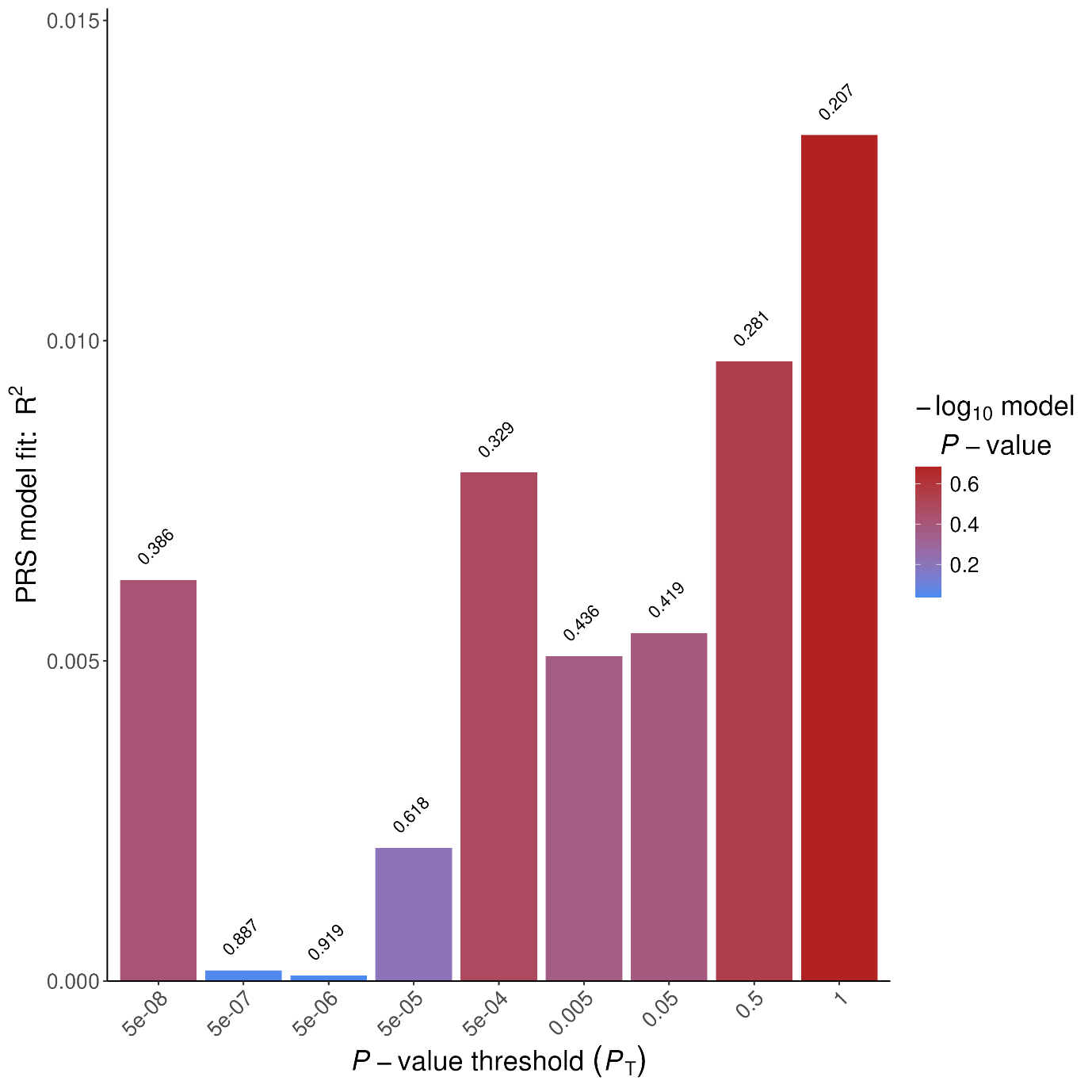

**Supplementary Figure S14: Prediction of Δ-9-THC-induced sadness by PRS of cannabis use disorder across nine p-value thresholds for SNP inclusion**

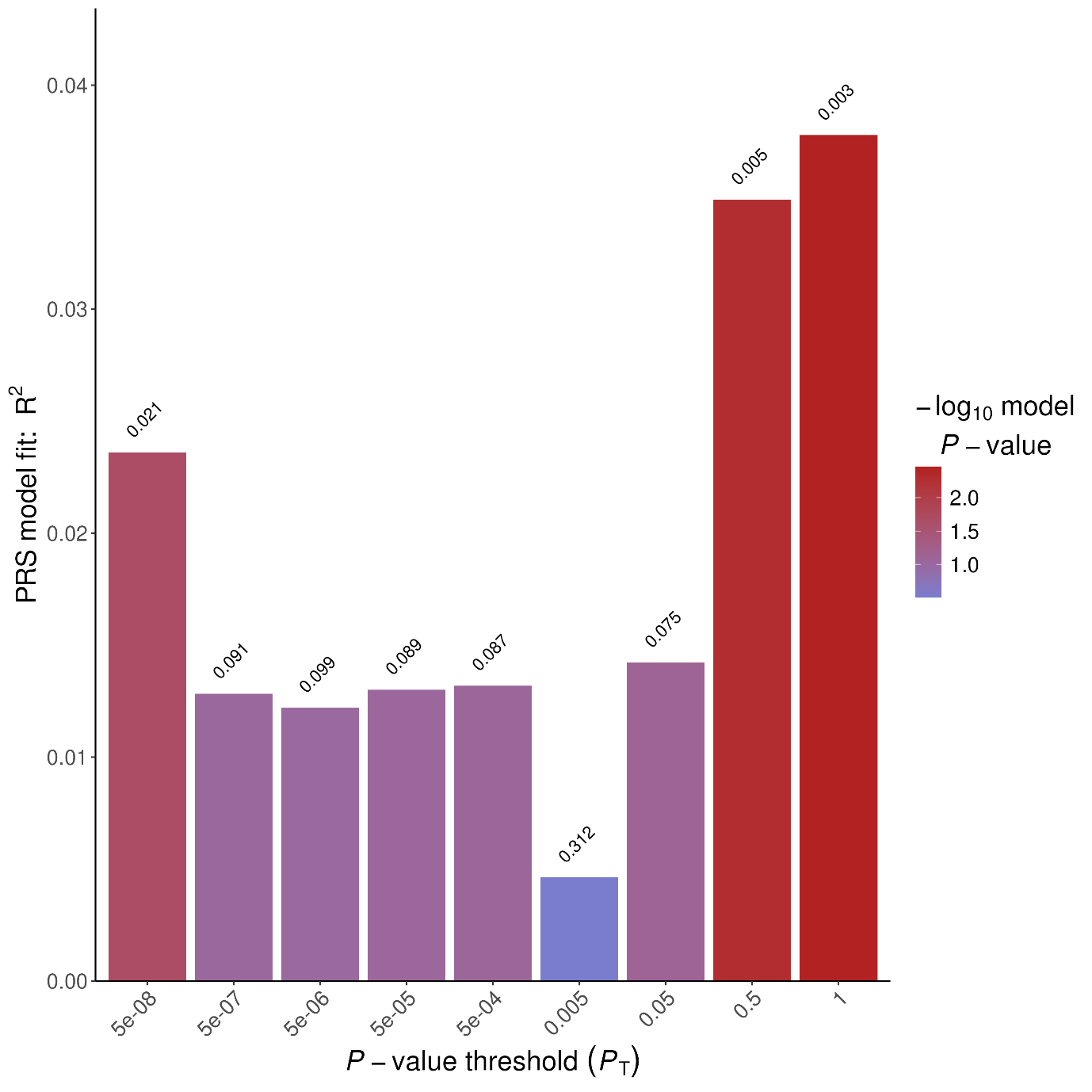

**Supplementary Figure S15: Prediction of Δ-9-THC-induced PANSS general symptoms by PRS of CYP3A4 expression across nine p-value thresholds for SNP inclusion**

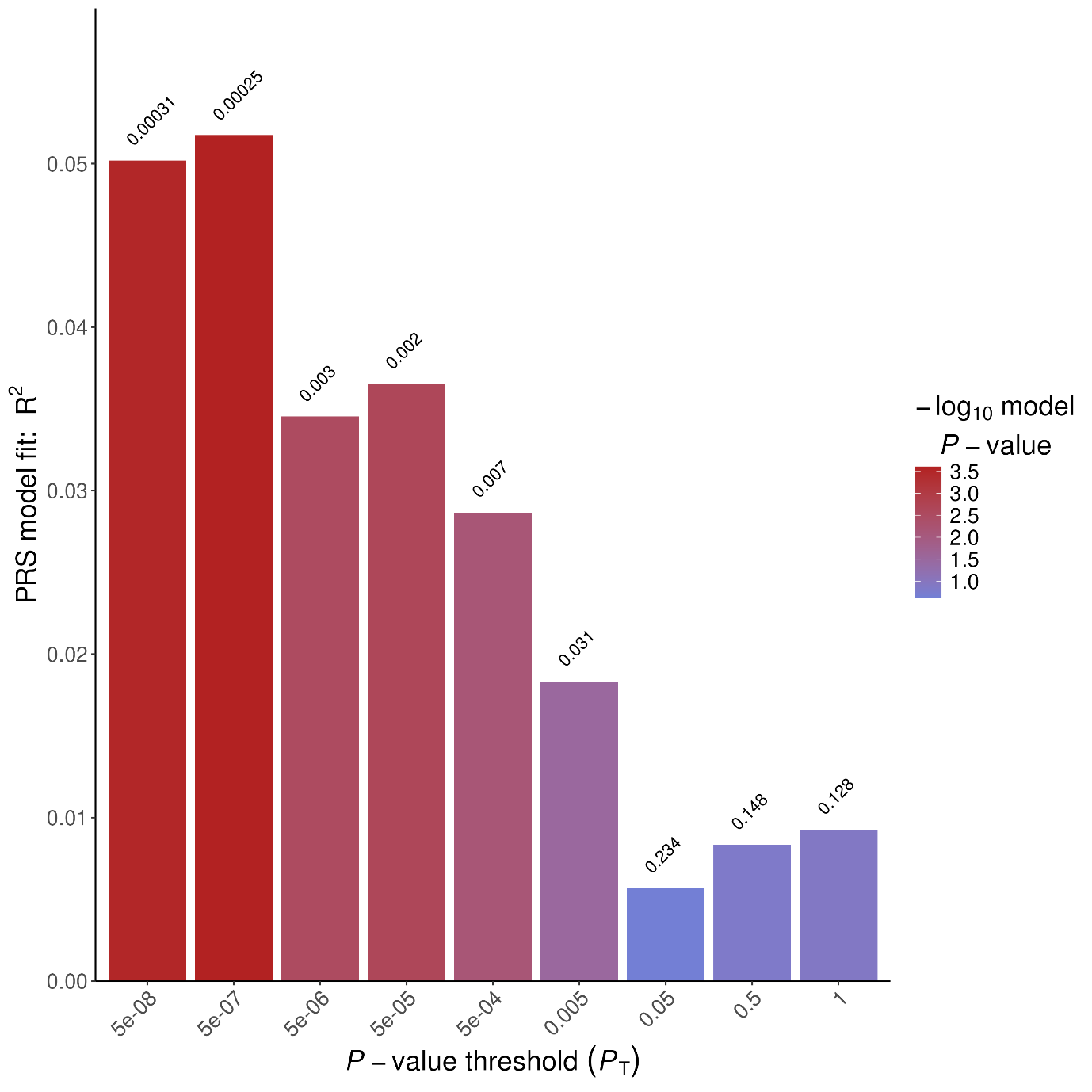

**Supplementary Figure S16: Prediction of Δ-9-THC-induced PANSS negative symptoms by PRS of CYP3A4 expression across nine p-value thresholds for SNP inclusion**

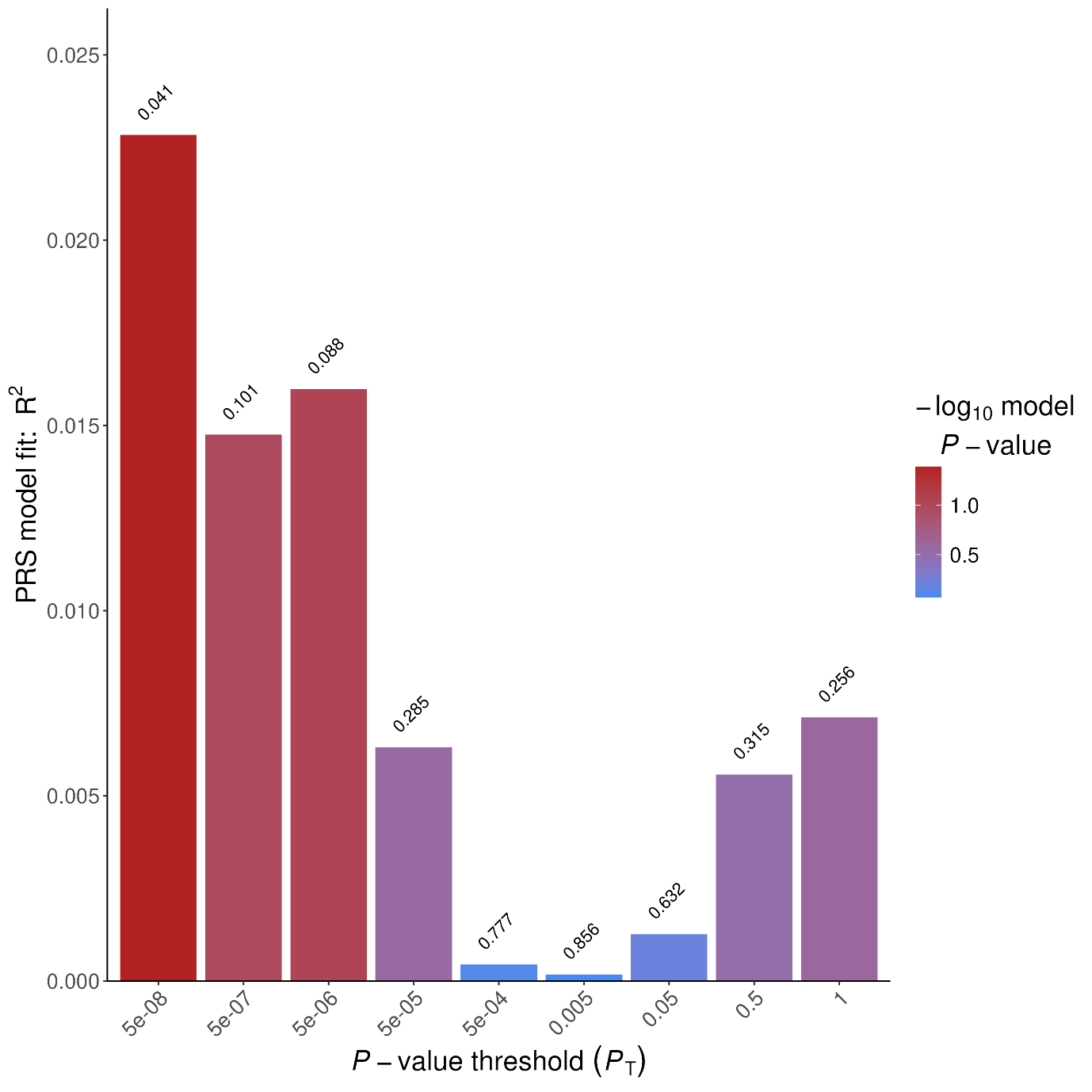

**Supplementary Figure S17: Prediction of Δ-9-THC-induced PANSS positive symptoms by PRS of CYP3A4 expression across nine p-value thresholds for SNP inclusion**

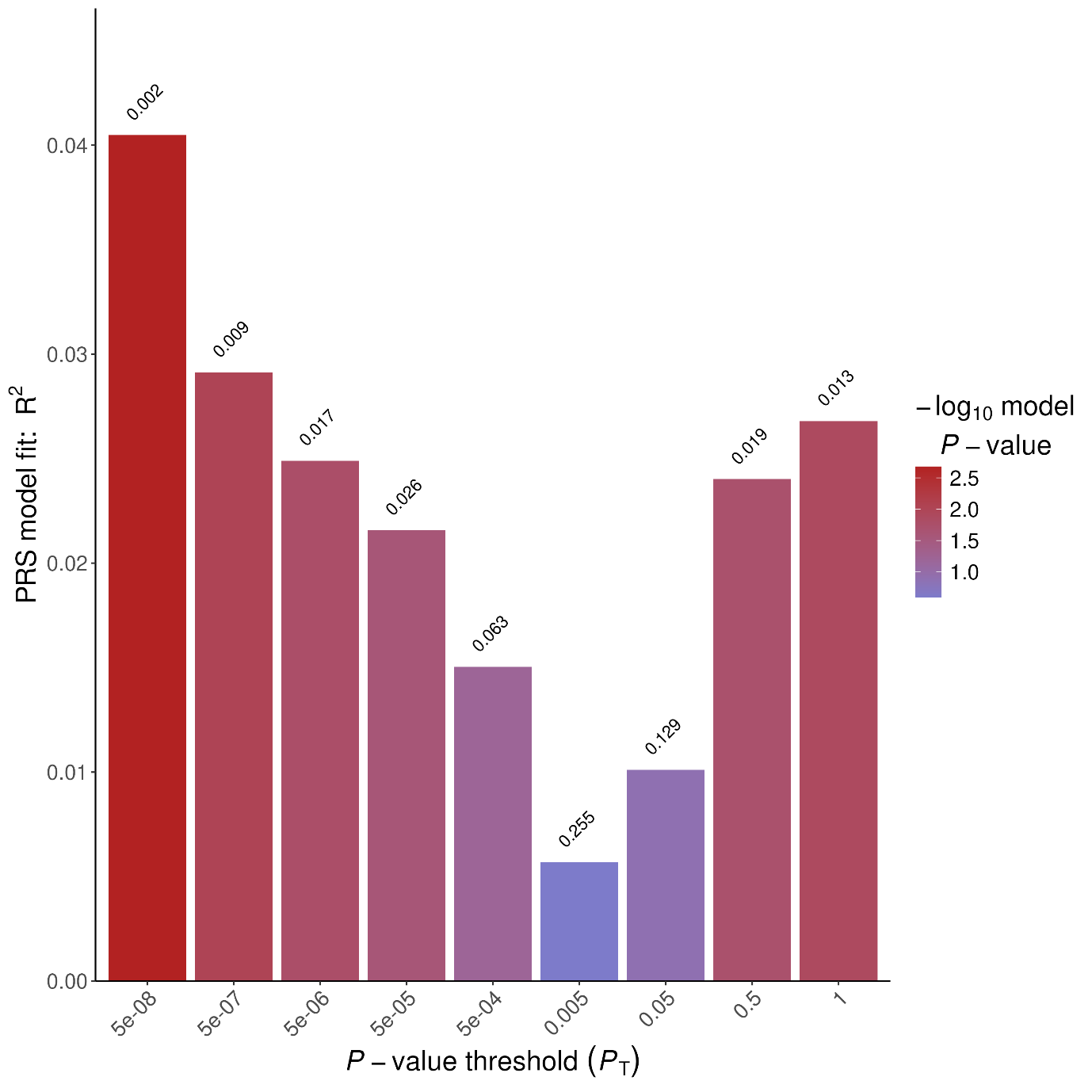

**Supplementary Figure S18: Prediction of Δ-9-THC-induced PANSS total score by PRS of CYP3A4 expression across nine p-value thresholds for SNP inclusion**

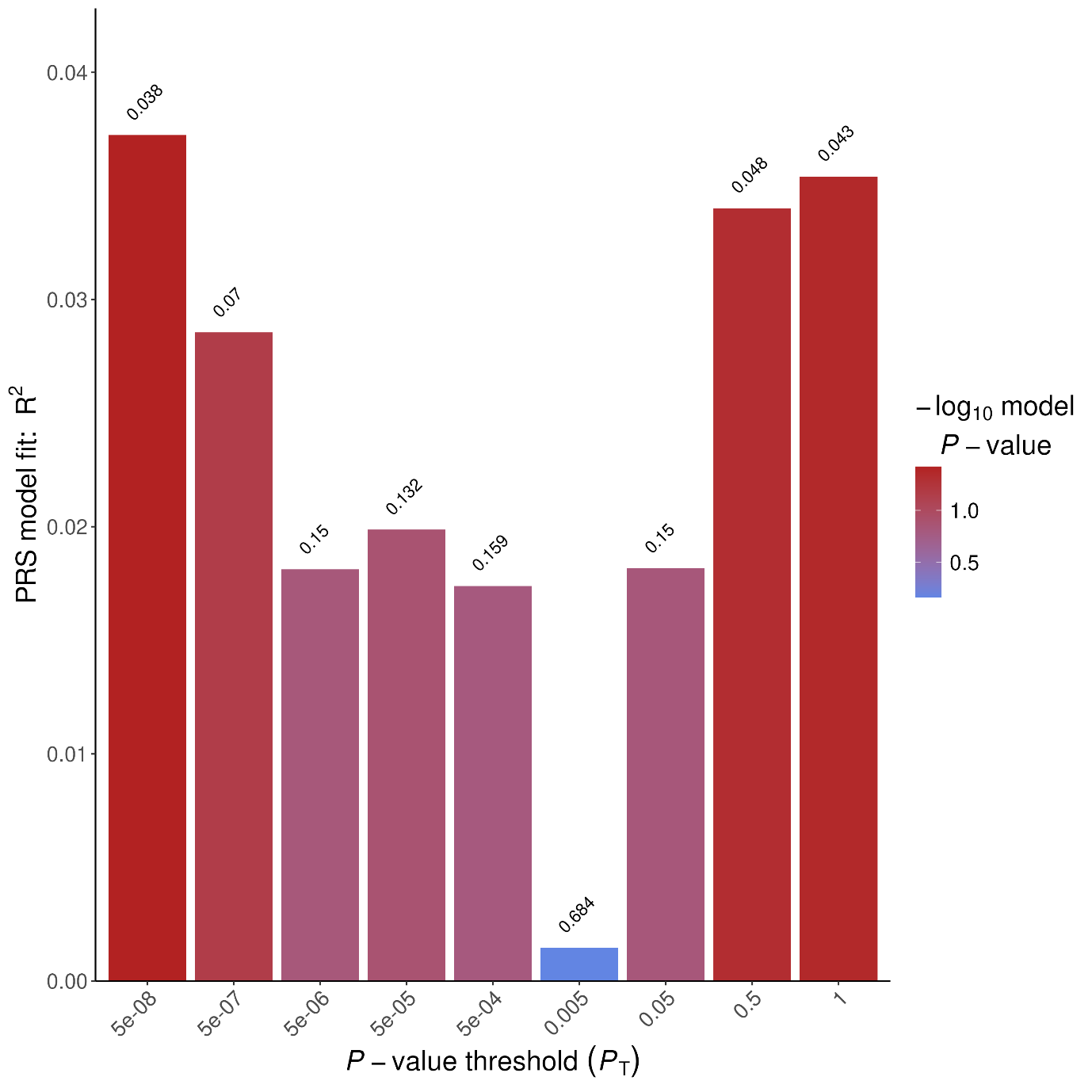

**Supplementary Figure S19: Prediction of Δ-9-THC-induced anxiety by PRS of CYP3A4 expression across nine p-value thresholds for SNP inclusion**

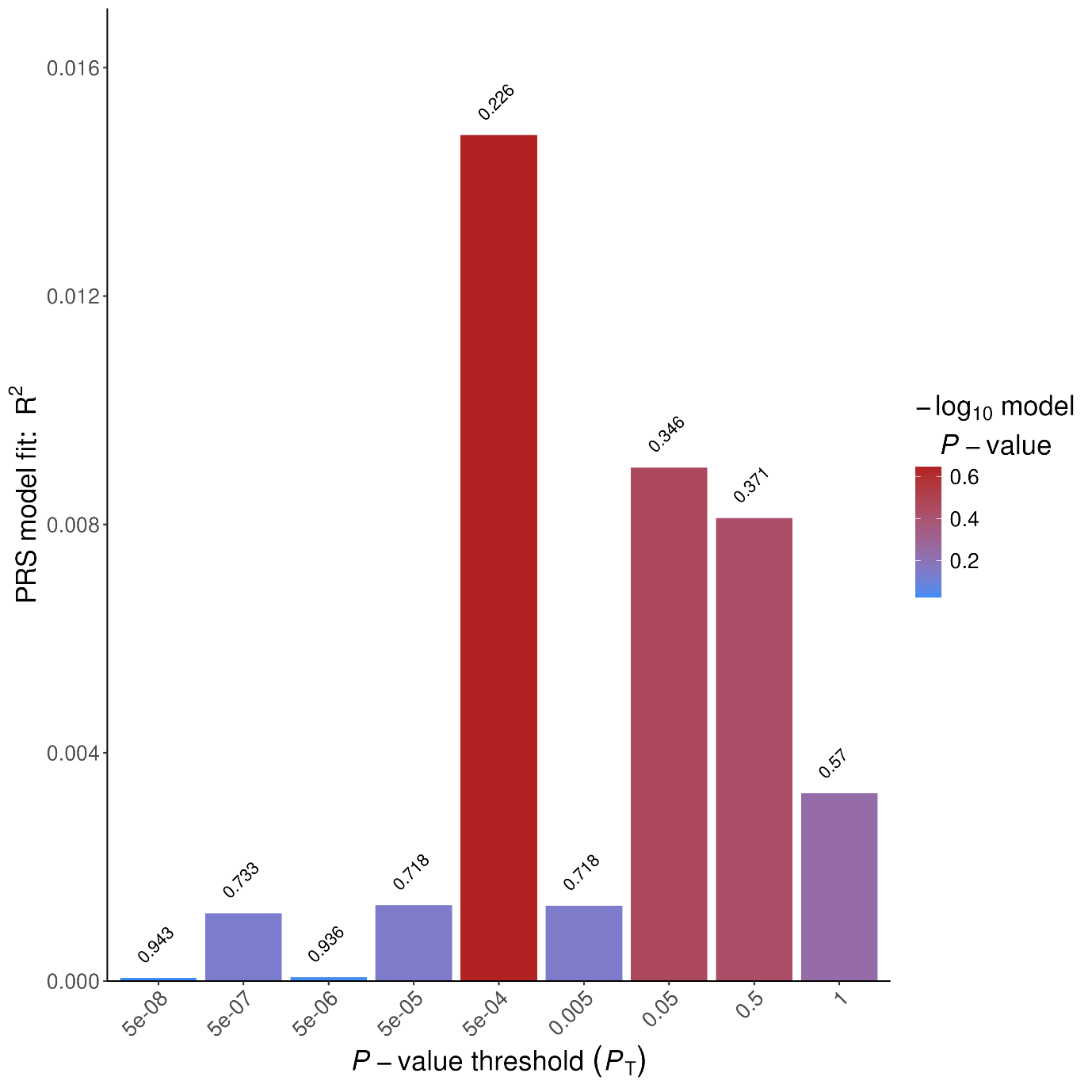

**Supplementary Figure S20: Prediction of Δ-9-THC-induced “high” by PRS of CYP3A4 expression across nine p-value thresholds for SNP inclusion**

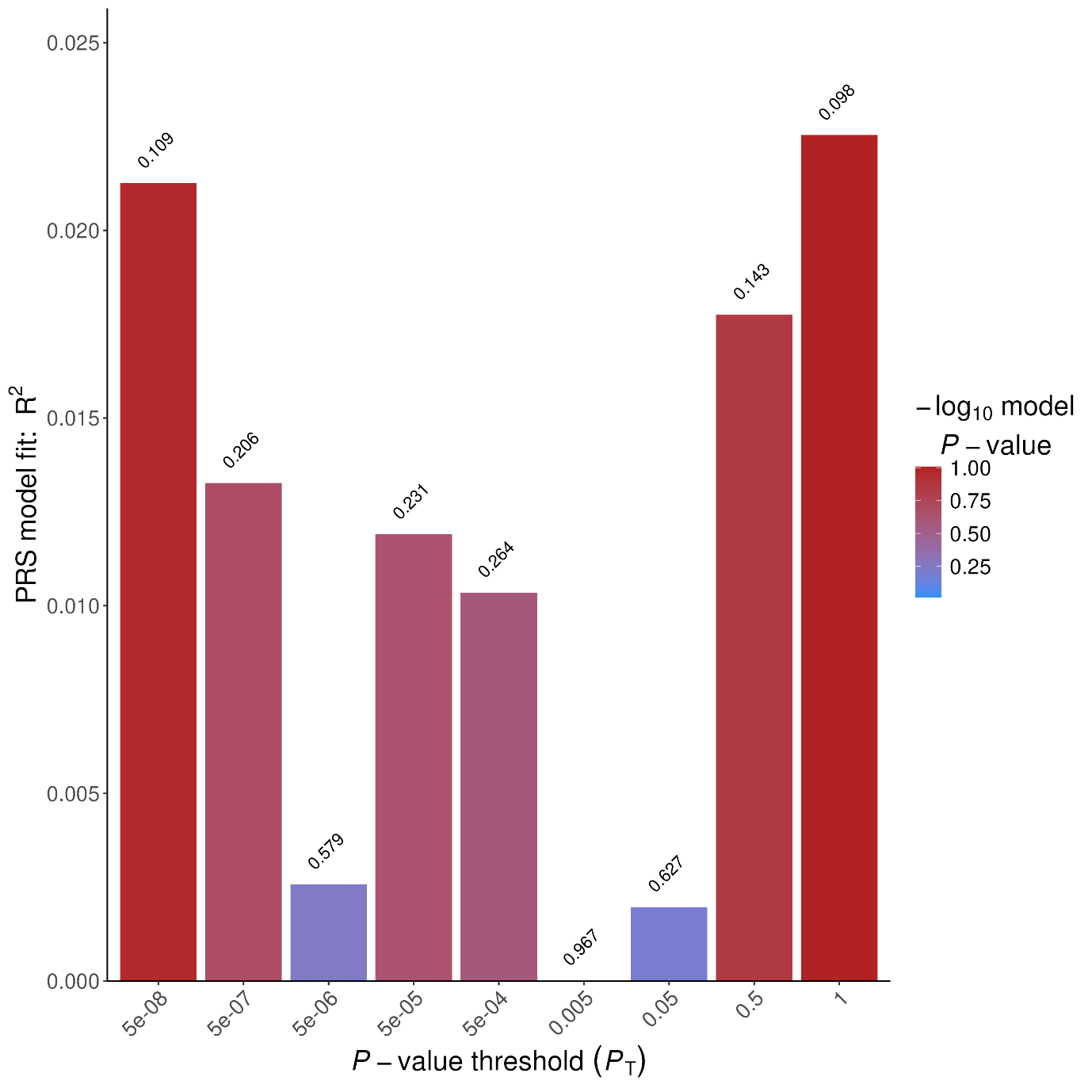

**Supplementary Figure S21: Prediction of Δ-9-THC-induced sadness by PRS of CYP3A4 expression across nine p-value thresholds for SNP inclusion**
